# Understanding priorities and needs for child and adolescent mental health in Greece from multiple informants: an open resource

**DOI:** 10.1101/2023.04.27.23288927

**Authors:** Anastasia Koumoula, Lauro Estivalete Marchionatti, Vasiliki Eirini Karagiorga, Julia Luiza Schafer, André Simioni, Arthur Caye, Aspasia Serdari, Konstantinos Kotsis, Maria Basta, Lilian Athanasopoulou, Vaios Dafoulis, Paraskevi Tsatsiopoulou, Nikos Zilikis, Evangelia Vergouli, Panagiota Balikou, Efi Kapsimali, Andromachi Mitropoulou, Alexandra Tzotzi, Nikanthi Klavdianou, Domna Zeleni, Sotiria Mitroulaki, Anna Botzaki, Giorgos Gerostergios, Giorgos Samiotakis, Giorgos Moschos, Ioanna Giannopoulou, Katerina Papanikolaou, Katerina Angeli, Nikolaos Scarmeas, Jill Emanuele, Kenneth Schuster, Eirini Karyotaki, Lily Kalikow, Katerina Pronoiti, Kathleen R. Merikangas, Peter Szatmari, Pim Cuijpers, Katholiki Georgiades, Michael P. Milham, Mimi Corcoran, Sarah Burke, Harold Koplewicz, Giovanni Abrahão Salum

## Abstract

**Background:** Evidence-based policymaking is needed so that health systems can address the gap in care for children and adolescents facing mental health challenges. We describe the development of an open-resource dataset providing a comprehensive assessment of the needs for child and adolescent mental health care in Greece.

**Methods:** This study is a part of the Child and Adolescent Mental Health Initiative (CAMHI), a program aiming to enhance mental health care capacity for children and adolescents across Greece. A comprehensive, mixed-method, community-based research was conducted in 2022/2023 to examine the current state, needs, barriers, and opportunities according to multiple viewpoints. Participants consisted of children, adolescents (including underrepresented minorities), caregivers, schoolteachers, and health professionals. We surveyed geographically distributed samples to assess mental health symptoms, mental health needs, literacy and stigma, service use and access, professional practices, training background, and training needs and preferences. Focus groups were conducted with informants to reach an in-depth understanding of those topics.

**Results:** We surveyed 1,756 caregivers, 1,201 children/adolescents, 404 schoolteachers, and 475 health professionals. Fourteen focus groups were conducted with the general and professional community. A repository with quantitative and qualitative findings informing multiple topics is now available for researchers, policymakers, and society [https://osf.io/crz6h/].

**Discussion:** This resource offers valuable data for assessing the needs and priorities for child and adolescent mental health care in Greece. It is now freely available to consult, and is expected to inform upcoming research and evidence-based professional training. This initiative may inspire similar ones in other countries, informing methodological strategies for researching mental health needs.

**Funding:** The Stavros Niarchos Foundation.

## 1. Introduction

Over the last decades, the mental healthcare system in Greece has been transitioning from a traditional in-patient treatment system to a community-oriented primary care model (Giannakopoulos and Anagnostopoulos 2016). Within this process, significant progress was achieved for child and adolescent mental health, including the establishment of specialized public services in a multi-sectoral system (Madianos 2020; G. Christodoulou et al. 2010). However, numerous challenges remain to be addressed. The financial crisis hindered funding, and affected policies destined for children and adolescents (N. G. Christodoulou and Anagnostopoulos 2013; Souliotis et al. 2017; Kolaitis and Giannakopoulos 2015). The distribution of resources is unequal, and there are deficits in availability of services and quality of care (N. G. Christodoulou and Anagnostopoulos 2013; Kolaitis and Giannakopoulos 2015; Petrea et al. 2020). There is a paucity of child and adolescent psychiatrists and other certified mental health professionals in the public sector, and gaps in professional training (N. G. Christodoulou and Kollias 2019; G. N. Christodoulou et al. 2012; Anargyros, Christodoulou, and Lappas 2021; Petrea et al. 2020). Moreover, there is a lack of information regarding the needs of children and adolescents, their families, and mental health professionals (Paleologou et al. 2018; Stierman et al. 2021).

To trace opportunities for strengthening community-based mental healthcare, it is necessary to have a comprehensive assessment of the mental health landscape in Greece. To achieve this, multiple methods can uncover complementary facets of mental health, reaching perspectives on necessities according to diverse viewpoints. Surveys are important sources to identify priorities that will inform resource allocation. In turn, qualitative assessment and community-based research may promote context-sensitive change, as it gives voice to the individuals on their necessities, attitudes, and dynamics related to mental health (Moser and Korstjens 2022). This bottom-up approach is of special relevance for children and adolescents, as they voice mental health challenges using non-clinical conceptions that often escape traditional assessments (Johns Hopkins Bloomberg School of Public Health and United Nations Children’s Fund 2022). Finally, for researching mental health services, the active engagement of professionals offer unique insights on real-world practices, standards of care, and workflows, which may inform tailored training programs and project capacity building.

For maximizing research impact, it is also necessary to have data and results freely accessible to both the scientific and general community. Addressing this issue, the open science movement advocates for scientific knowledge to be openly shared, as well as for the construction of collaborative frameworks for building and disseminating research material (Tennant et al. 2016). This intends to optimize scientific efforts, amplifying its dissemination and thus increasing possible applications. We recently reviewed publications on prevalence estimates, assessment tools, and intervention trials published in Greece (Koumoula et al. 2022) and found that most of the datasets were not freely available, hindering progress and reuse of data to answer relevant questions to child mental health in Greece.

The Child and Adolescent Mental Health Initiative (CAMHI) is a program aiming to enhance child and adolescent mental health care capacity and to help strengthen the infrastructure for the prevention, assessment, and treatment of mental health struggles faced by children and adolescents across Greece. As part of this initiative, we performed a comprehensive assessment of the current state and needs in this field from the perspective of multiple viewpoints, including scores of mental health symptoms, mental health literacy and stigma, service use and access, professional practices, training background, and training needs and preferences. A mixed-method research strategy was employed, with qualitative focus groups and quantitative surveys gathering multiple information from children, adolescents, caregivers, mental health professionals, and teachers. Here, we describe this open-resource dataset, which is available for use by scientists, policymakers, and the general community.

## 2. Methods

We report the development of a repository following a convergent design, in which quantitative and qualitative data are concurrently collected aiming at combining their results to obtain a more complete understanding of the topic (Creswell and Clark 2017). Research participants were key informants from the general and professional community involved in the welfare and health care systems, namely children and adolescents, their families, mental health care professionals, pediatricians, teachers, and NGO members. Written informed consent was obtained from all participants and the research was approved by the Research Ethics Committee of the Democritus University of Thrace [approval number: ΔΠΘ/ΕΗΔΕ/42772/307]. Survey data was collected and preserved according to the General Data Protection Regulation (GDPR) National Policy (European Parliament and The Council 2016), being handled unidentified and kept under a password-protected system with access restricted to research members for a one-year period. Relevant material and records concerning the development of this research can be accessed on our webpage located on the Open Science Framework [https://osf.io/crz6h/], a platform for managing and storing shared knowledge projects (Foster and Deardorff 2017).

In the quantitative section, geographically distributed samples of the informant groups were assessed between September 2022 and January 2023 through a cross-sectional survey composed of validated instruments and questions covering areas of interest (see below for detailed recruitment strategies). The survey with children, adolescents, and caregivers assessed: (a) frequency of mental health problems; (b) mental health needs; (c) mental health literacy and stigma; (d) mental health services and access to care. The survey with teachers and healthcare professionals assessed: (1) mental health literacy and stigma; (2) professional practices and experience; (3) training background; (4) training needs and perspectives. For this arm of the research, we followed the study design items of the Strengthening the Reporting of Observational Studies in Epidemiology (STROBE) (refer to **Supplementary Table 1** for checklist) (von Elm et al. 2014).

As for the qualitative section, focus groups were conducted to reach an in-depth comprehension on the target topics and generate insight on solutions with the active participation of community members. To ensure the representation of vulnerable populations, some groups were composed of members of LGBTQIA+ groups, refugees, or ethnic minorities. Groups were focused on assessing: (a) views on wellness, mental health, and mental health problems; (b) mental health stigma and discrimination; (c) barriers, facilitators, and opportunities for mental health care; (d) mental health services and access. For teachers and health professionals, discussions also focused on (e) training needs and perspectives. For the items pertaining to this arm of the project, we followed the items on data collection and design from the Consolidated Criteria for Reporting Qualitative Research (COREQ) (refer to **Supplementary Table 2** for the checklist) (Tong, Sainsbury, and Craig 2007).

### 2.1. Surveys

#### 2.1.1. Measures

**Table 1** shows the selected instruments and **Table 2** displays the scope of developed questions that were used in the survey, whilst **Figure 1** and **Figure 2** depict how they were applied to each sample. The initial part of the survey consisted of general socio-demographic questions collecting relevant data from all respondents, including preliminary questions on mental health for children, adolescents, and caregivers (whether they face mental health problems, have a diagnosis, medication use, and professional assistance). Then, for each domain of inquiry, we consulted the literature to select locally validated instruments that assess relevant constructs (see **Supplementary Table 3** for their characteristics and psychometrics). If no validated Greek version was available, we performed a cross-cultural adaptation of a selected instrument following a five-stage validated procedure (see **Supplementary Table 4** for an outline of the procedure; detailed registers of the adaptation are available at [https://osf.io/crz6h/]) (Beaton et al. 2000). For a few topics (training background, training needs and perspectives, professional practice and experience, service use and access, and mental health needs), we could not locate adequate instruments covering sufficient aspects of interest, so we developed a set of questions (see **Table 2** for their scope). In this procedure, a team of local and international experts within our research group formulated survey items considering the context of participants, which were then selected and refined after extensive discussion until reaching a final set of questions. The full version of the questionnaire is available at [https://osf.io/crz6h/].

**Figure 1.**
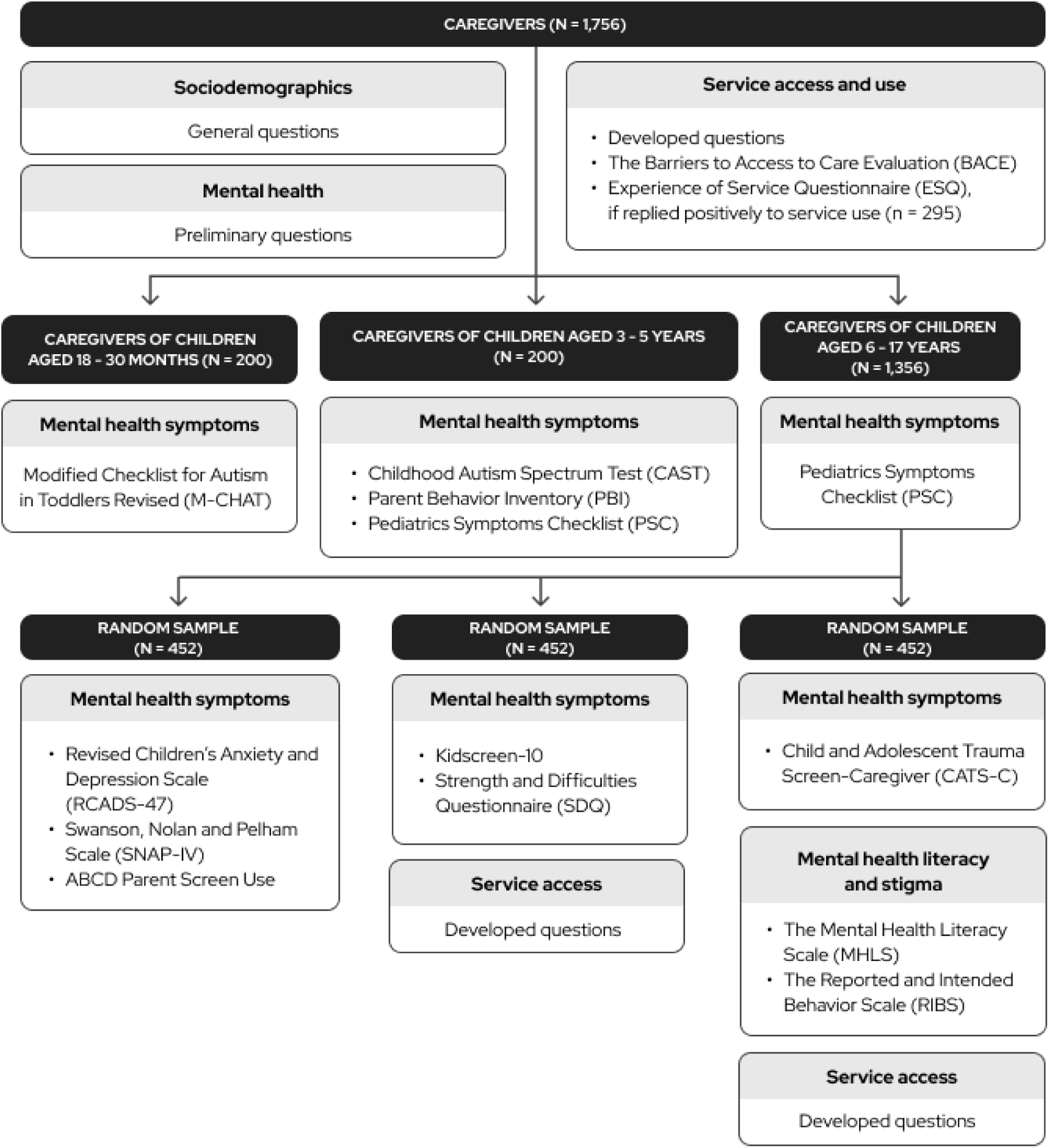
Survey design: participants and instruments (caregivers)

**Figure 2.**
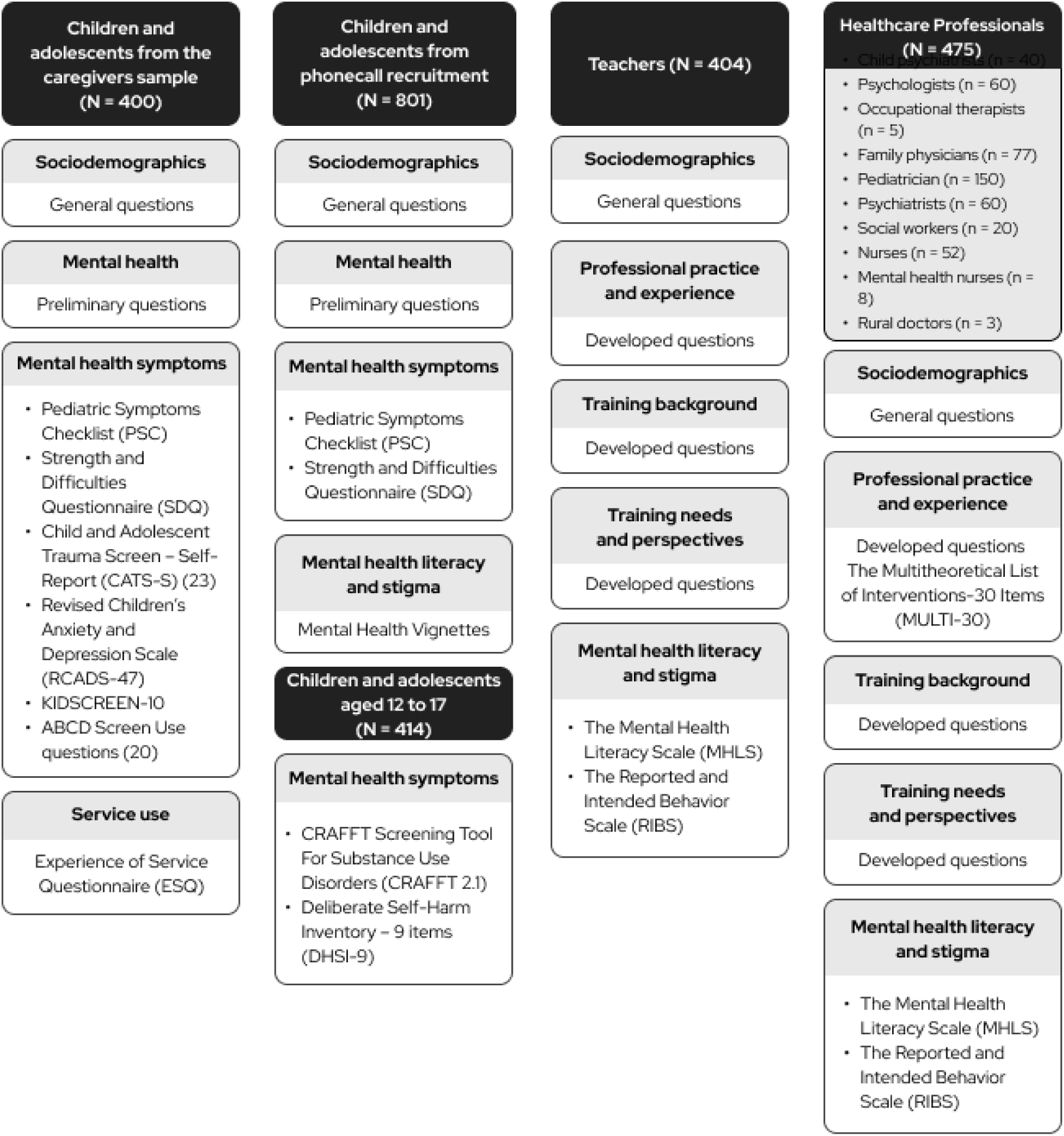
Survey design: participants and instruments (children, adolescents, health professionals and teachers)

**Table 1.**
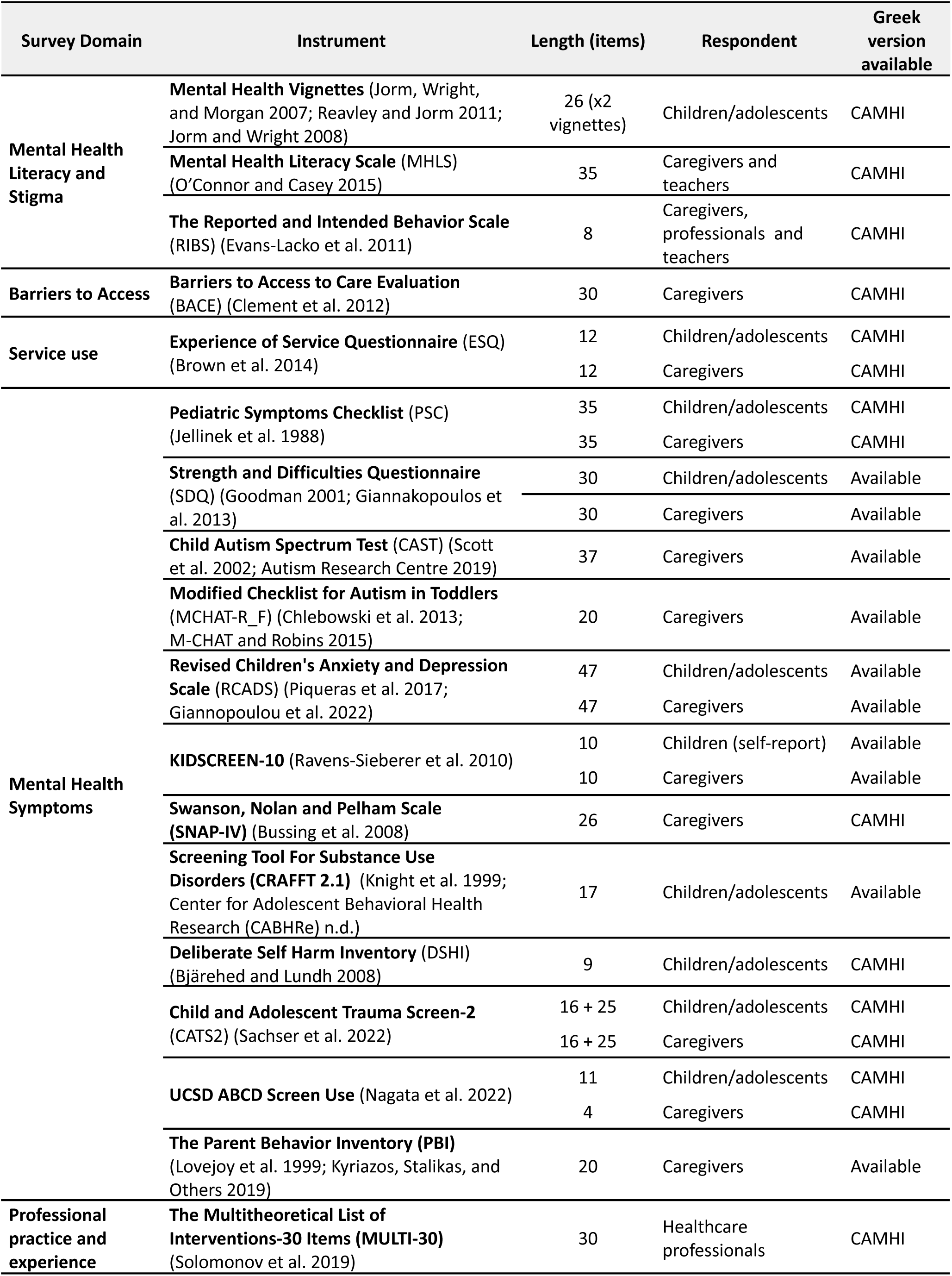
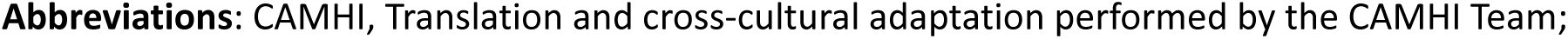
Survey Instruments.

| Survey Domain | Instrument | Length (items) | Respondent | Greek version available |
| --- | --- | --- | --- | --- |
| **Mental Health Literacy and Stigma** | **Mental Health Vignettes** (Jorm, Wright, and Morgan 2007; Reavley and Jorm 2011; Jorm and Wright 2008) | 26 (x2 vignettes) | Children/adolescents | CAMHI |
|  | **Mental Health Literacy Scale (MHLS)** (O'Connor and Casey 2015) | 35 | Caregivers and teachers | CAMHI |
|  | **The Reported and Intended Behavior Scale (RIBS)** (Evans-Lacko et al. 2011) | 8 | Caregivers, professionals and teachers | CAMHI |
| **Barriers to Access** | **Barriers to Access to Care Evaluation (BACE)** (Clement et al. 2012) | 30 | Caregivers | CAMHI |
| **Service use** | **Experience of Service Questionnaire (ESQ)** (Brown et al. 2014) | 12 | Children/adolescents | CAMHI |
|  |  | 12 | Caregivers | CAMHI |
| **Mental Health Symptoms** | **Pediatric Symptoms Checklist (PSC)** (Jellinek et al. 1988) | 35 | Children/adolescents | CAMHI |
|  |  | 35 | Caregivers | CAMHI |
|  | **Strength and Difficulties Questionnaire (SDQ)** (Goodman 2001; Giannakopoulos et al. 2013) | 30 | Children/adolescents | Available |
|  |  | 30 | Caregivers | Available |
|  | **Child Autism Spectrum Test (CAST)** (Scott et al. 2002; Autism Research Centre 2019) | 37 | Caregivers | Available |
|  | **Modified Checklist for Autism in Toddlers (MCHAT-R_F)** (Chlebowski et al. 2013; M-CHAT and Robins 2015) | 20 | Caregivers | Available |
|  | **Revised Children's Anxiety and Depression Scale (RCADS)** (Piqueras et al. 2017; Giannopoulou et al. 2022) | 47 | Children/adolescents | Available |
|  |  | 47 | Caregivers | Available |
|  | **KIDSCREEN-10** (Ravens-Sieberer et al. 2010) | 10 | Children (self-report) | Available |
|  |  | 10 | Caregivers | Available |
|  | **Swanson, Nolan and Pelham Scale (SNAP-IV)** (Bussing et al. 2008) | 26 | Caregivers | CAMHI |
|  | **Screening Tool For Substance Use Disorders (CRAFT 2.1)** (Knight et al. 1999; Center for Adolescent Behavioral Health Research (CABHRe) n.d.) | 17 | Children/adolescents | Available |
|  | **Deliberate Self Harm Inventory (DSHI)** (Bjärehed and Lundh 2008) | 9 | Children/adolescents | CAMHI |
|  | **Child and Adolescent Trauma Screen-2 (CATS2)** (Sachser et al. 2022) | 16 + 25 | Children/adolescents | CAMHI |
|  |  | 16 + 25 | Caregivers | CAMHI |
|  | **UCSD ABCD Screen Use** (Nagata et al. 2022) | 11 | Children/adolescents | CAMHI |
|  |  | 4 | Caregivers | CAMHI |
|  | **The Parent Behavior Inventory (PBI)** (Lovejoy et al. 1999; Kyriazos, Stalikas, and Others 2019) | 20 | Caregivers | Available |
| **Professional practice and experience** | **The Multitheoretical List of Interventions-30 Items (MULTI-30)** (Solomonov et al. 2019) | 30 | Healthcare professionals | CAMHI |

**Abbreviations:** CAMHI, Translation and cross-cultural adaptation performed by the CAMHI Team;

**Table 2.**
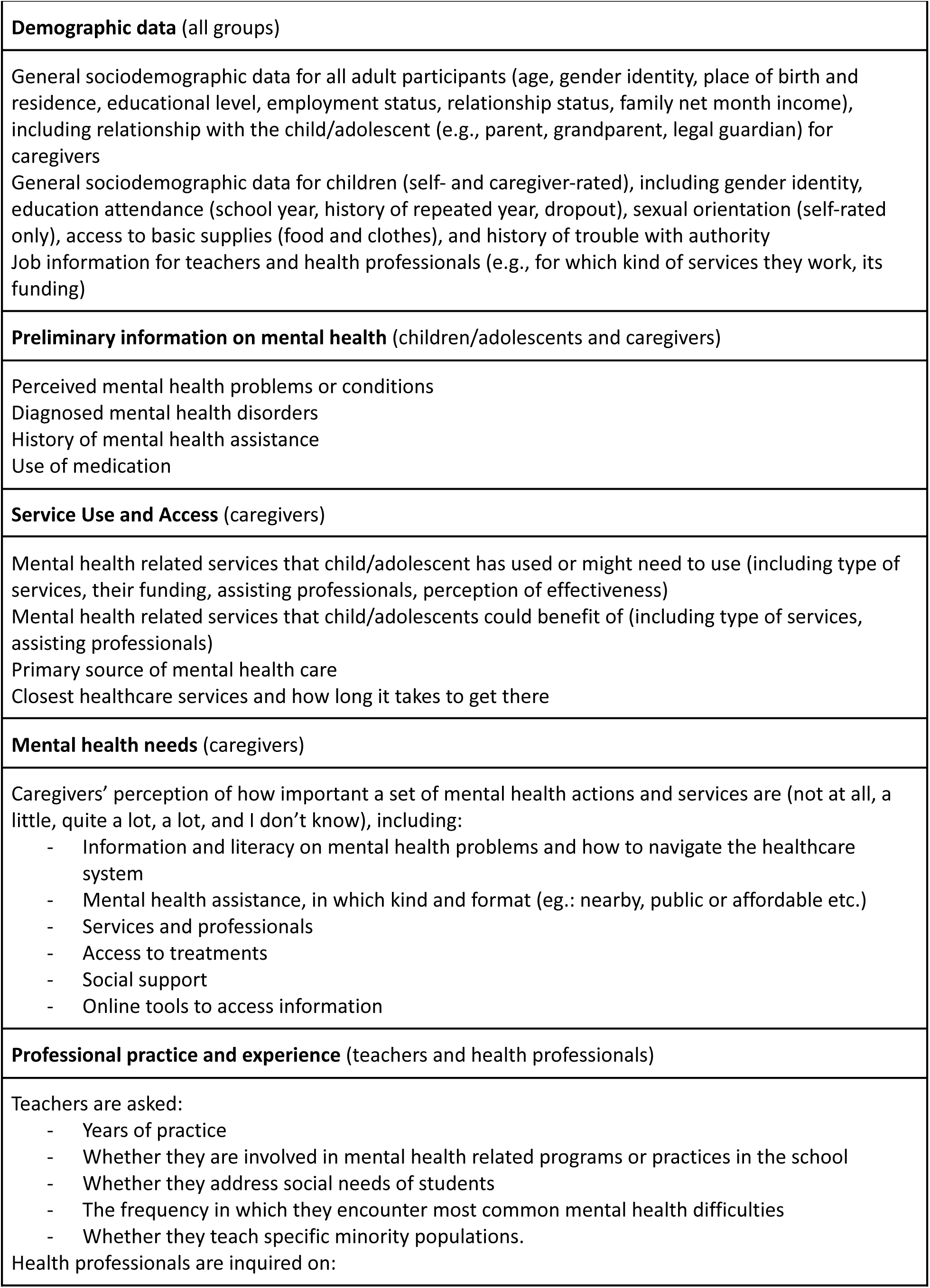
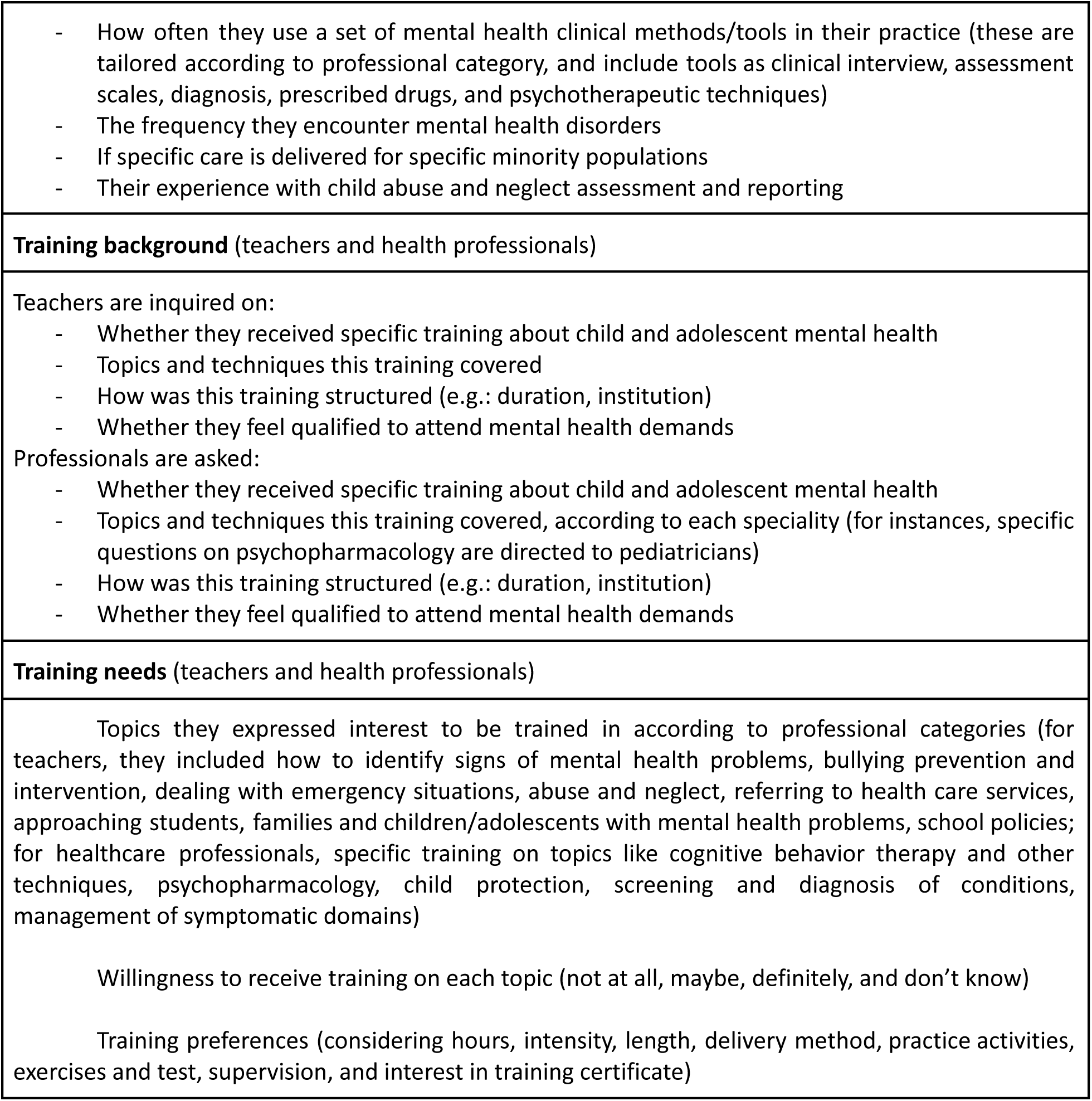
Scope of questions developed to survey research topics.

|  |
| --- |
| **Demographic data** (all groups) |
| General sociodemographic data for all adult participants (age, gender identity, place of birth and residence, educational level, employment status, relationship status, family net month income), including relationship with the child/adolescent (e.g., parent, grandparent, legal guardian) for caregivers<br>General sociodemographic data for children (self- and caregiver-rated), including gender identity, education attendance (school year, history of repeated year, dropout), sexual orientation (self-rated only), access to basic supplies (food and clothes), and history of trouble with authority<br>Job information for teachers and health professionals (e.g., for which kind of services they work, its funding) |
| **Preliminary information on mental health** (children/adolescents and caregivers) |
| Perceived mental health problems or conditions<br>Diagnosed mental health disorders<br>History of mental health assistance<br>Use of medication |
| **Service Use and Access** (caregivers) |
| Mental health related services that child/adolescent has used or might need to use (including type of services, their funding, assisting professionals, perception of effectiveness)<br>Mental health related services that child/adolescents could benefit of (including type of services, assisting professionals)<br>Primary source of mental health care<br>Closest healthcare services and how long it takes to get there |
| **Mental health needs** (caregivers) |
| Caregivers' perception of how important a set of mental health actions and services are (not at all, a little, quite a lot, a lot, and I don't know), including:<br>- Information and literacy on mental health problems and how to navigate the healthcare system<br>- Mental health assistance, in which kind and format (eg.: nearby, public or affordable etc.)<br>- Services and professionals<br>- Access to treatments<br>- Social support<br>- Online tools to access information |
| **Professional practice and experience** (teachers and health professionals) |
| Teachers are asked:<br>- Years of practice<br>- Whether they are involved in mental health related programs or practices in the school<br>- Whether they address social needs of students<br>- The frequency in which they encounter most common mental health difficulties<br>- Whether they teach specific minority populations.<br>Health professionals are inquired on: |

#### 2.1.2. Recruitment and participants

##### Survey with Caregivers

A nationwide sample of caregivers was recruited to a self-applied online questionnaire according to regional and offspring gender and age quotas following the census distribution (see **Supplementary Table 5** for quotas) (Hellenic Statistical Authority 2012). The questionnaire consisted of questions and instruments on service access and use, literacy and stigma, parenting practices, and mental health symptoms of general and specific conditions (see **Figure 1** for details of which instruments were applied to the sample and its subsets). Participants were reached through a proprietary online respondent panel (Kantar Group and Affiliates n.d.). This panel was developed based on a census frame and its members were rigorously profiled using over 500 data points. Recruitment occurred online via social media and website campaigns, search engine optimization (SEO), panelists’ friends referrals, and affiliate networks. To avoid self-selection, surveys were automatically routed to respondents based on an algorithm that combines representativeness (via random allocation), appropriate frequency of sampling for each panelist, and management of sample quotas if required. For each survey, a pre-specified maximum number of 1.400 participants applies. Panelists who have been profiled as parents or legal guardians of children under 18 years old were invited to this survey following the algorithm set to manage regional location quotas. A screening question was applied to ensure that the children/adolescents they cared for were between 1.5 and 17 years old at intake.

##### Survey with Children and Adolescents

Surveys were conducted with two groups of children and adolescents answering different sets of instruments (see **Figure 2** for details). The first group consisted of the offspring of the surveyed caregivers, who were invited to a self-applied online questionnaire with instruments measuring mental health symptoms to match equivalent instruments rated by their caregivers. They provided measures on service use and mental health symptoms (including general psychopathology, screen use, and anxiety and depression). A second group was selected by random landline phone calls that recruited children and adolescents aged 8 to 17 years old across the Greek territory (maximum of one per household) to a telephone survey, following census quotas on region, gender, and two age groups (see **Supplementary Table 5** for quotas) (Hellenic Statistical Authority 2012). An automatic dialer system generated landline and mobile phone numbers to make automatic calls after removing non-operational numbers. This group answered general psychopathology measures, as well as mental health stigma and discrimination questionnaires based on the mental health vignettes (further described in the qualitative arm). Adolescents aged 12 to 17 years old in this second group also filled instruments on self-harm behavior and substance use disorder, as well as questions on gender and sexuality.

##### Survey with Teachers

A list of 274 regular and special education schools were randomly sampled from the five key areas where the CAMHI hubs are located (Athens, Thessaloniki, Ioannina, Alexandroupoli, and Heraklion). Schoolteachers were recruited in person at the place of work, and then phone call interviews were held at the respondents convenience with a scheduled appointment. To ensure socio-demographic representativeness, the samples were proportionally distributed across and within districts according to the population size following census distribution (see **Supplementary Table 5** for quotas) (Hellenic Statistical Authority 2012). Teachers answered questionnaires on demographics, mental health literacy and stigma, professional experience, training skills, and perspective on training needs (see **Figure 2**).

##### Survey with Healthcare Professionals

Healthcare professionals from different fields of practice (child psychiatrists, psychologists, occupational therapists, family physicians, pediatricians, psychiatrists, social workers, and nurses) were randomly sampled from a proprietary database (the IQVIA OneKey) containing contact and profiling information on the specialties intended for the survey (IQVIA n.d.). Sampling was done randomly within each specialty area, respecting geographical distribution quotas following the database profiling across the five hub areas designated (see **Supplementary Table 5** for quotas). Participants were called and invited to respond to questionnaires that were adapted according to speciality (**Figure 2**). Interviews were held online with a computer-assisted interview program (CAWI), being guided by an IQVIA executive by phone to maximize completion rate and quality.

#### 2.1.3. Data analysis

Descriptive analysis was performed to provide measures of central tendency, frequency, and dispersion for each instrument or subset of questions. As relevant, analysis was stratified by subgroups. Statistical analysis was performed in the software R version 3.6.2 (R Core Team 2013).

### 2.2. Focus groups

We conducted 14 focus groups (10 online and 4 in person) of 60 to 115 minutes duration, each composed by 8 to 10 members of an informant group (see **Table 3** for a detailed description). Participants were recruited from the five cities where the CAMHI hubs are located after an initial online search for schools, parents’ associations, health centers, professional associations, LGBTQIA+ organizations, and legal channels. We contacted these institutions to obtain a list of potential participants who were then invited to participate in the groups via a telephone call. Pomak and Roma adolescents were recruited through a cultural mediator involved in the community. All participants received a 100-euro reward as compensation for their time. Details of the recruitment process according to each group can be seen in **Supplementary Table 6**.

**Table 3.** Description of focus groups.

| Population | Number of participants | Location | Duration |
| --- | --- | --- | --- |
| **General population** |  |  |  |
| Adolescents aged 15 years-old | 10 | Online | 90 min |
| Caregivers of 8 years-old children | 10 | Online | 115 min |
| Caregivers of 15 years-old adolescents | 11 | Online | 115 min |
| Children aged 8 years-old | 9 | Athens | 60 min |
| **Underrepresented minorities** |  |  |  |
| LGBTQIA+ adolescents | 8 | Athens | 90 min |
| Roma adolescents | 10 | Athens | 90 min |
| Pomak adolescents | 10 | Alexandroupoli | 90 min |
| Refugee unaccompanied minors | 5 | Athens | 90 min |
| **Professional community** |  |  |  |
| NGO leaders | 9 | Online | 115 min |
| Psychologists and social workers | 9 | Online | 115 min |
| Child psychiatrists, family physicians, and pediatricians | 10 | Online | 115 min |
| Nurses and nurse assistants | 10 | Online | 115 min |
| Speech therapists & occupational therapists | 10 | Online | 115 min |
| School teachers | 10 | Online | 115 min |

A vignette-based discussion was carried out to illustrate children and adolescents facing mental health disorders and prompt impressions from participants (see **Supplementary Table 7** for complete vignettes). To represent both internalizing and externalizing prevalent conditions, the children’s vignettes featured cases of social anxiety and attention-deficit/hyperactivity disorder (ADHD), while adolescents’ vignettes featured depression and conduct disorder. The vignettes on social anxiety and depression were adapted to Greek based on the Australian National Survey on Youth Mental Health Literacy, a work assessing beliefs towards mental health (Jorm, Wright, and Morgan 2007). The vignettes on conduct disorder and ADHD were constructed by our research team following a similar process. All vignettes were adapted according to the cultural background of each focus group by using a common name in that language.

Each focus group was guided by a local moderator with experience in the field and the assistance of manuals containing anchor questions (refer to **Supplementary Table 8** for the structure of the manuals and to the Open Science Framework webpage [https://osf.io/crz6h/] for its complete version; see **Supplementary Table 6** for moderators’ credentials). Researchers had no previous relationship with the participants, and disclosed they were professional scholars conducting a work for deepening the understanding on the research topics. The manuals were constructed by an international panel of specialists within our research team in a brainstorming process that generated candidate questions that were then selected, discussed, and refined to reach a final version of anchor questions for the manual. To guarantee that the manuals were context-sensitive, a multicultural panel of specialists with local and external expertise participated in the process, including professionals of diverse fields of activity and experience with refugees and other vulnerable groups. Manuals were adapted according to necessities and specificities of each focus group.

The focus groups’ sessions were video recorded, resulting in a written transcript in the spoken language of the group (Greek or Farsi) and a translated version to English. Written notes were taken in real-time as the group occurred. Immediately following each focus group, the research team conducted a debrief session with the moderator to gather top-of-mind perceptions, using the heightened awareness of the moment to generate initial ideas for analysis and insights to guide future focus group sessions. Considering data was translated to English to be made available for analysis, transcripts were not returned to participants for comment or correction. As a first processing of the transcripts, we summarized the discussion contained within each focus group, providing a narrative description of the major points elicited in the sessions and its supporting quotations.

To illustrate the content of the focus groups, the research team (LEM, EV) looked for pertinent discussions about mental health concepts and stigma, which were purposefully sampled for a brief report. This was examined by attending general principles of inductive thematic analysis, and coding was performed by one researcher (LEM) and discussed in regular meetings with other members of the team (JLF, GAS, EV) (Braun and Clarke 2006). For each focus group, we looked at the selected data to detect meaningful patterns in responses. Initial codes were generated and then clustered into organizing central concepts to delineate broad findings. We then compared those findings across the different groups, usefully arranging under four overarching populations that most accurately nested results: professionals, caregivers, children/adolescents of general population, and children/adolescents of underrepresented minorities (namely, Pomak, Roma, LGBTQIA+, and refugee adolescents). The findings were then outlined as bullet points in the form of sentences that more succinctly described their attributes.

## 3. Results

### 3.1) Survey

Between 2022 and 2023, we invited 8,863 caregivers, 954 children/adolescents related to these caregivers, 19,008 children/adolescents from random telephone calls, 409 teachers, and 1,700 healthcare professionals across a range of specialists to participate in the survey. Following no response, decline to participation, or dropout during questionnaire, a total number of 1,756 caregivers (response rate 19,81%), 400 children/adolescents related to caregivers (response rate 41,92%), 801 children from random telephone call recruitment (response rate 4,74%), 404 teachers (response rate 99,02%), and 475 healthcare professionals (response rate 27,94%) completed the questionnaires. The demographics and relevant characteristics of each group are presented in **Table 4**. Preliminary descriptive measures for screening instruments on general psychopathology, as rated by caregivers and children/adolescents, are presented in **Table 5**. For navigating across the complete results of the dataset, we also developed an online dashboard which is freely available in [https://rpubs.com/camhi/sdashboard]. Raw data sheets will be made available at Open Science Framework webpage [https://osf.io/crz6h/] one year after the completion of data collection, and specific variables can be already requested by researchers.

**Table 4.**
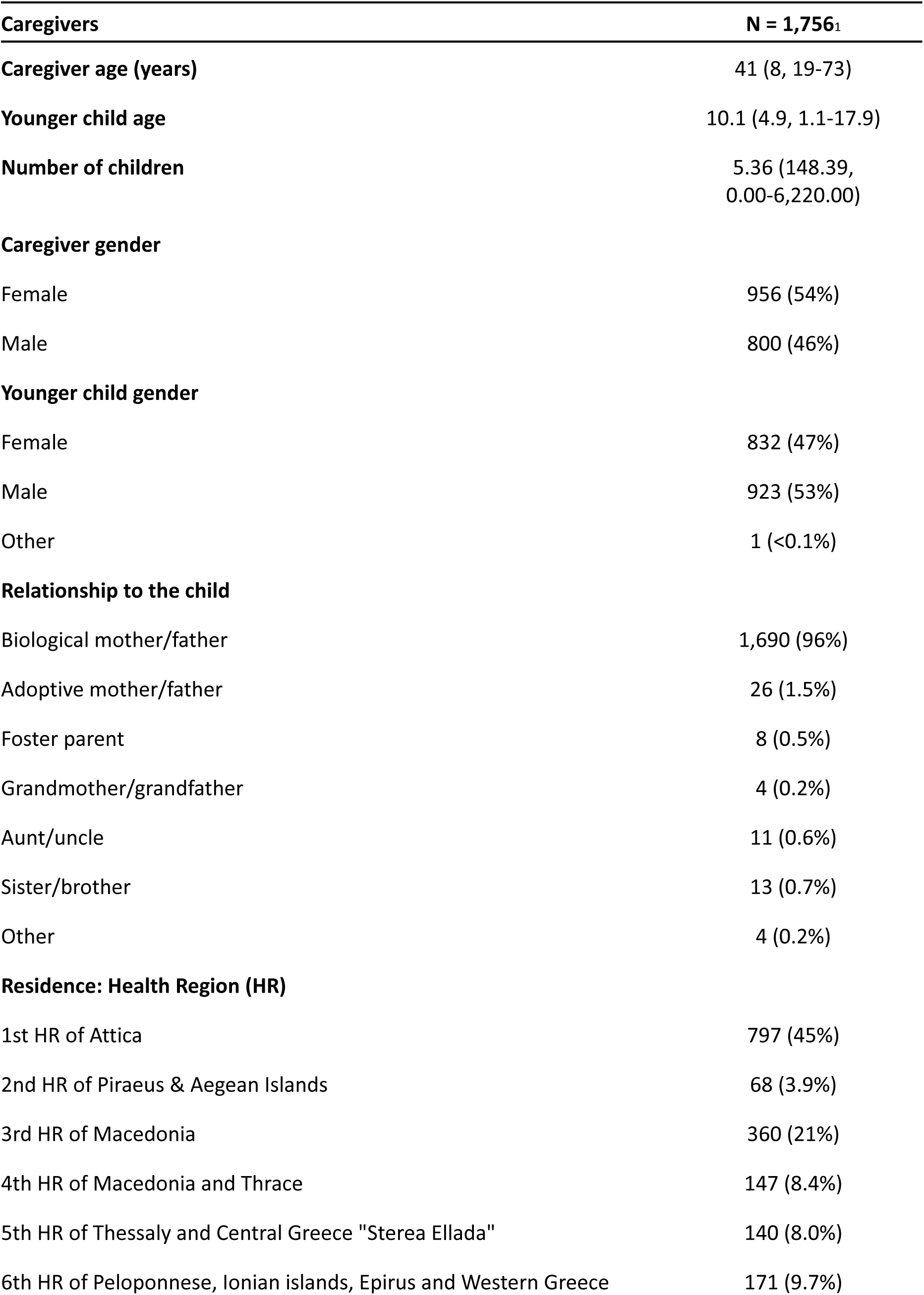
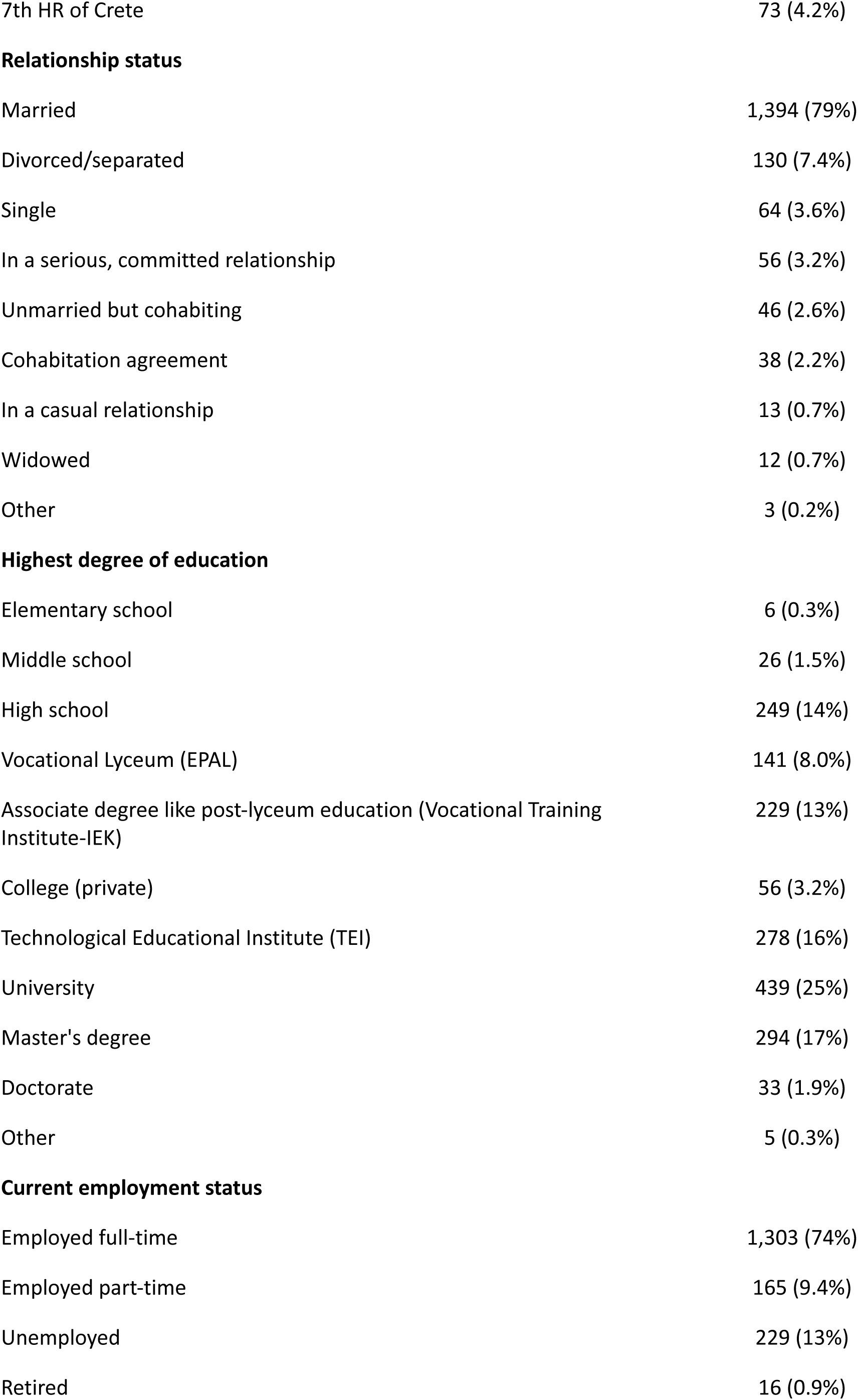
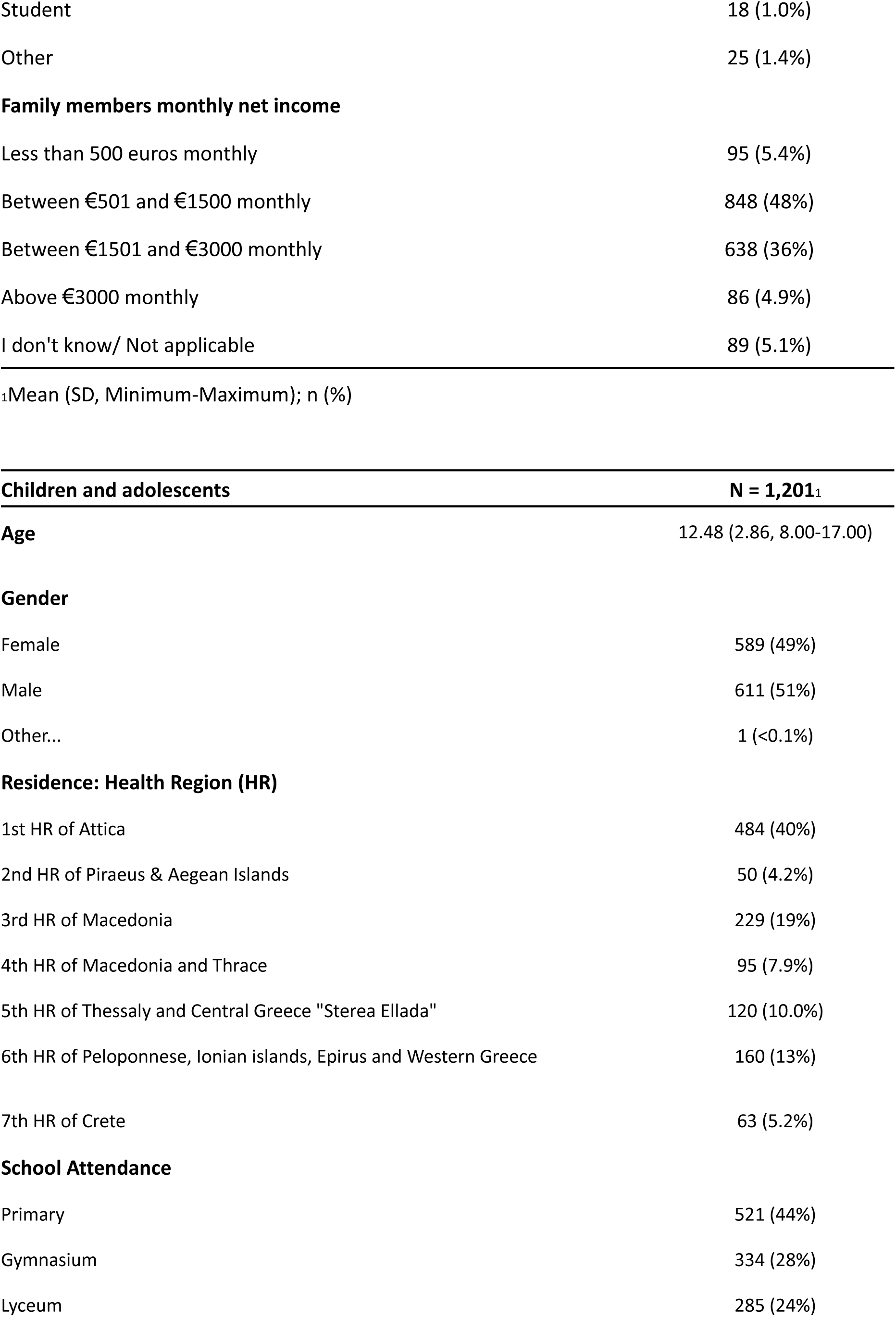
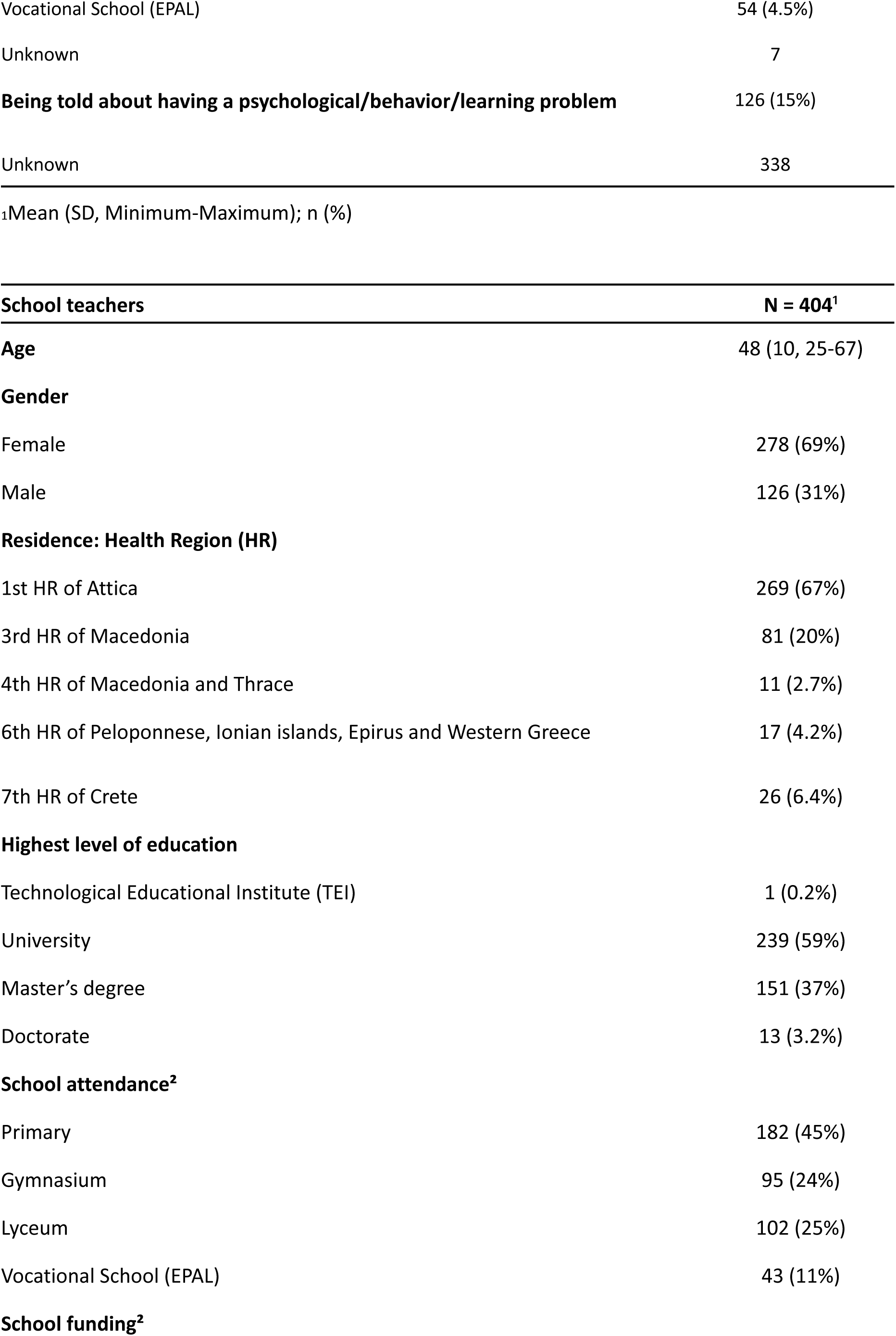
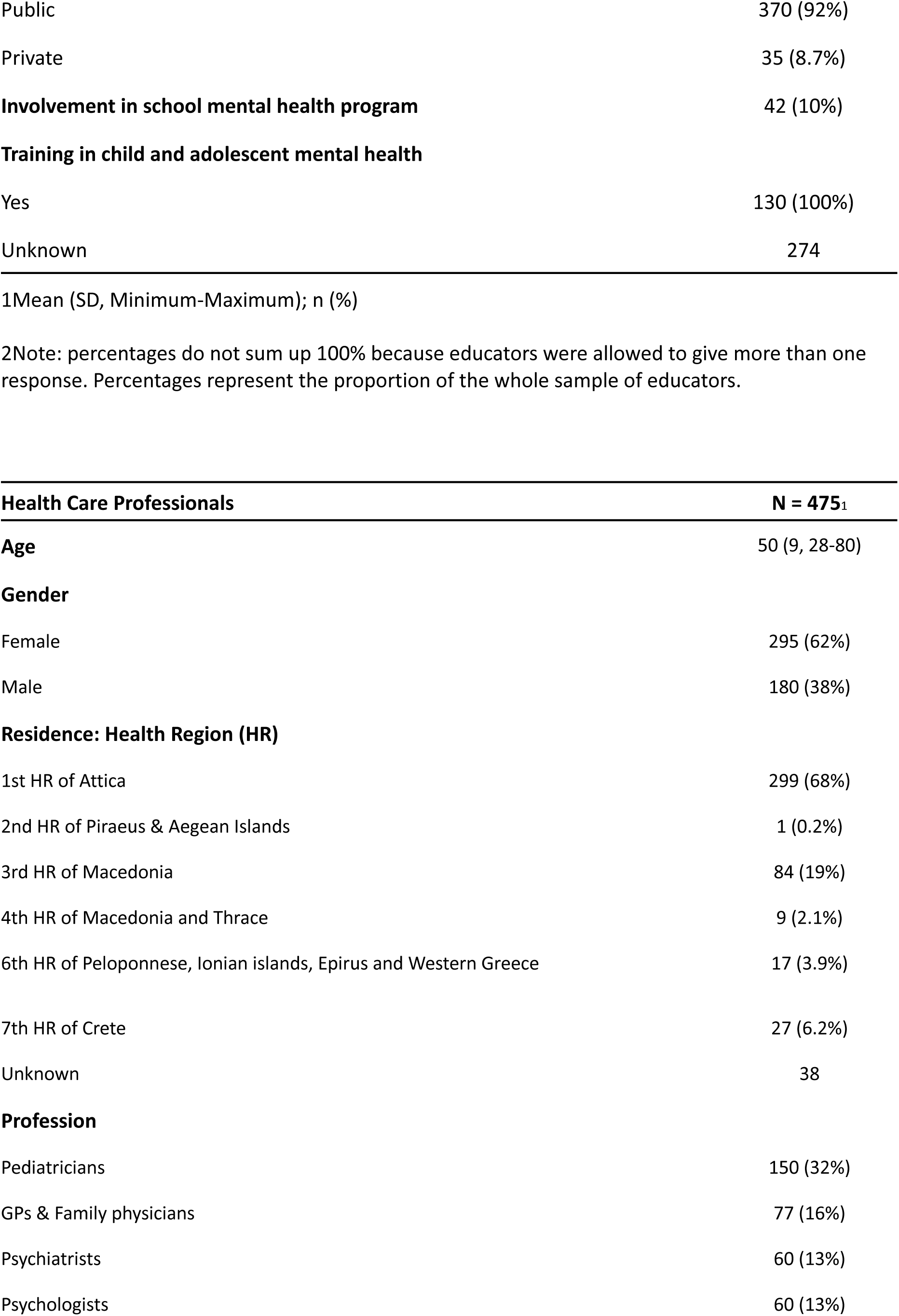
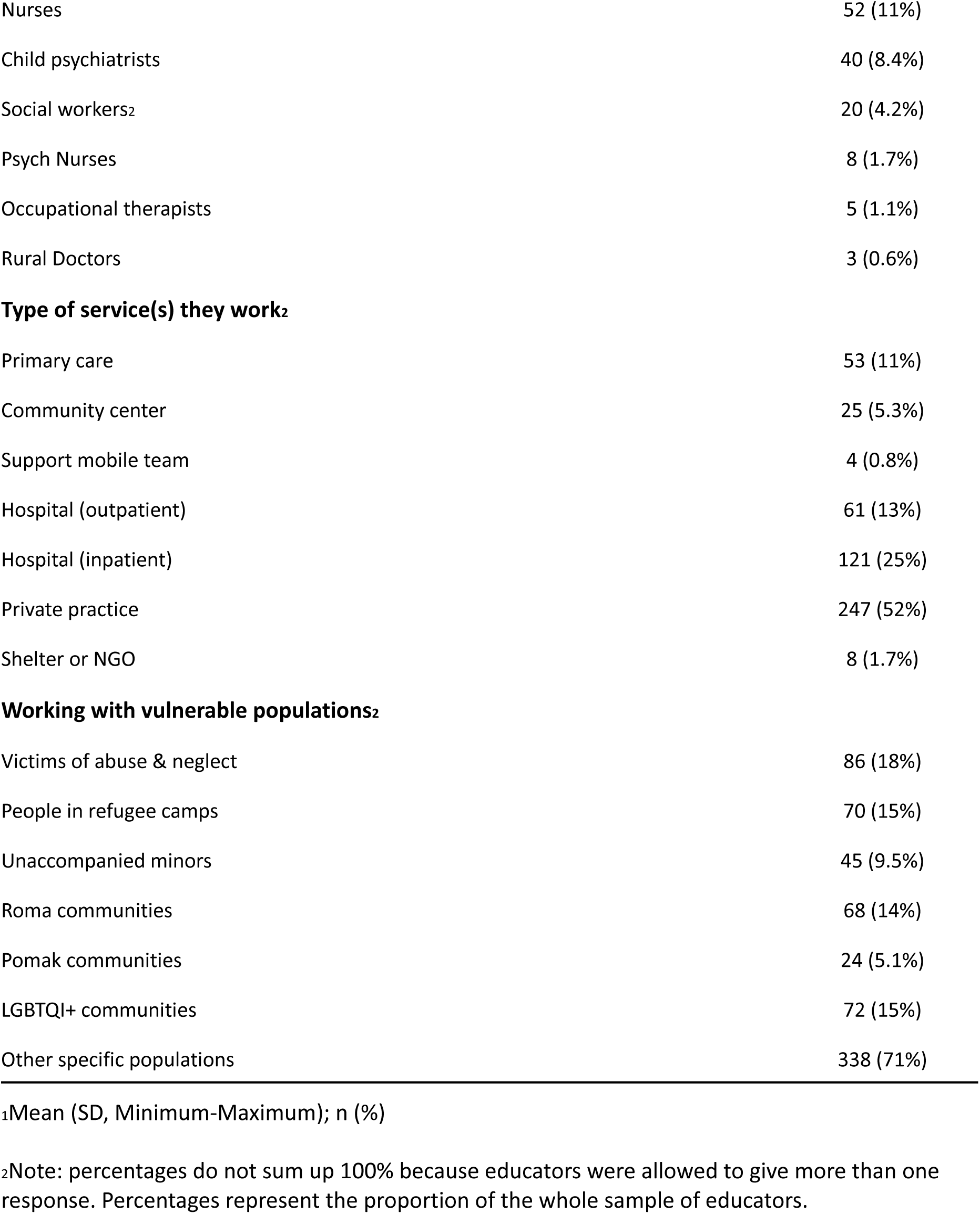
Sociodemographic Characteristics of Survey Participants.

| **Caregivers** | **N = 1,756<sub>1</sub>** |
| --- | --- |
| **Caregiver age (years)** | 41 (8, 19-73) |
| **Younger child age** | 10.1 (4.9, 1.1-17.9) |
| **Number of children** | 5.36 (148.39,<br>0.00-6,220.00) |
| **Caregiver gender** |  |
| Female | 956 (54%) |
| Male | 800 (46%) |
| **Younger child gender** |  |
| Female | 832 (47%) |
| Male | 923 (53%) |
| Other | 1 (<0.1%) |
| **Relationship to the child** |  |
| Biological mother/father | 1,690 (96%) |
| Adoptive mother/father | 26 (1.5%) |
| Foster parent | 8 (0.5%) |
| Grandmother/grandfather | 4 (0.2%) |
| Aunt/uncle | 11 (0.6%) |
| Sister/brother | 13 (0.7%) |
| Other | 4 (0.2%) |
| **Residence: Health Region (HR)** |  |
| 1st HR of Attica | 797 (45%) |
| 2nd HR of Piraeus & Aegean Islands | 68 (3.9%) |
| 3rd HR of Macedonia | 360 (21%) |
| 4th HR of Macedonia and Thrace | 147 (8.4%) |
| 5th HR of Thessaly and Central Greece "Sterea Ellada" | 140 (8.0%) |
| 6th HR of Peloponnese, Ionian islands, Epirus and Western Greece | 171 (9.7%) |
| 7th HR of Crete | 73 (4.2%) |
| **Relationship status** |  |
| Married | 1,394 (79%) |
| Divorced/separated | 130 (7.4%) |
| Single | 64 (3.6%) |
| In a serious, committed relationship | 56 (3.2%) |
| Unmarried but cohabiting | 46 (2.6%) |
| Cohabitation agreement | 38 (2.2%) |
| In a casual relationship | 13 (0.7%) |
| Widowed | 12 (0.7%) |
| Other | 3 (0.2%) |
| **Highest degree of education** |  |
| Elementary school | 6 (0.3%) |
| Middle school | 26 (1.5%) |
| High school | 249 (14%) |
| Vocational Lyceum (EPAL) | 141 (8.0%) |
| Associate degree like post-lyceum education (Vocational Training Institute-IEK) | 229 (13%) |
| College (private) | 56 (3.2%) |
| Technological Educational Institute (TEI) | 278 (16%) |
| University | 439 (25%) |
| Master's degree | 294 (17%) |
| Doctorate | 33 (1.9%) |
| Other | 5 (0.3%) |
| **Current employment status** |  |
| Employed full-time | 1,303 (74%) |
| Employed part-time | 165 (9.4%) |
| Unemployed | 229 (13%) |
| Retired | 16 (0.9%) |
| Student | 18 (1.0%) |
| Other | 25 (1.4%) |
| **Family members monthly net income** |  |
| Less than 500 euros monthly | 95 (5.4%) |
| Between €501 and €1500 monthly | 848 (48%) |
| Between €1501 and €3000 monthly | 638 (36%) |
| Above €3000 monthly | 86 (4.9%) |
| I don't know/ Not applicable | 89 (5.1%) |
| 1Mean (SD, Minimum-Maximum); n (%) |  |
| **Children and adolescents** | **N = 1,201<sup>1</sup>** |
| **Age** | 12.48 (2.86, 8.00-17.00) |
| **Gender** |  |
| Female | 589 (49%) |
| Male | 611 (51%) |
| Other... | 1 (<0.1%) |
| **Residence: Health Region (HR)** |  |
| 1st HR of Attica | 484 (40%) |
| 2nd HR of Piraeus & Aegean Islands | 50 (4.2%) |
| 3rd HR of Macedonia | 229 (19%) |
| 4th HR of Macedonia and Thrace | 95 (7.9%) |
| 5th HR of Thessaly and Central Greece "Sterea Ellada" | 120 (10.0%) |
| 6th HR of Peloponnese, Ionian islands, Epirus and Western Greece | 160 (13%) |
| 7th HR of Crete | 63 (5.2%) |
| **School Attendance** |  |
| Primary | 521 (44%) |
| Gymnasium | 334 (28%) |
| Lyceum | 285 (24%) |
| Vocational School (EPAL) | 54 (4.5%) |
| Unknown | 7 |
| **Being told about having a psychological/behavior/learning problem** | 126 (15%) |
| Unknown | 338 |
| 1Mean (SD, Minimum-Maximum); n (%) |  |
| **School teachers** | **N = 404<sup>1</sup>** |
| **Age** | 48 (10, 25-67) |
| **Gender** |  |
| Female | 278 (69%) |
| Male | 126 (31%) |
| **Residence: Health Region (HR)** |  |
| 1st HR of Attica | 269 (67%) |
| 3rd HR of Macedonia | 81 (20%) |
| 4th HR of Macedonia and Thrace | 11 (2.7%) |
| 6th HR of Peloponnese, Ionian islands, Epirus and Western Greece | 17 (4.2%) |
| 7th HR of Crete | 26 (6.4%) |
| **Highest level of education** |  |
| Technological Educational Institute (TEI) | 1 (0.2%) |
| University | 239 (59%) |
| Master's degree | 151 (37%) |
| Doctorate | 13 (3.2%) |
| **School attendance<sup>2</sup>** |  |
| Primary | 182 (45%) |
| Gymnasium | 95 (24%) |
| Lyceum | 102 (25%) |
| Vocational School (EPAL) | 43 (11%) |
| **School funding<sup>2</sup>** |  |
| Public | 370 (92%) |
| Private | 35 (8.7%) |
| **Involvement in school mental health program** | 42 (10%) |
| **Training in child and adolescent mental health** |  |
| Yes | 130 (100%) |
| Unknown | 274 |

| **Health Care Professionals** | **N = 475<sub>1</sub>** |
| --- | --- |
| **Age** | 50 (9, 28-80) |
| **Gender** |  |
| Female | 295 (62%) |
| Male | 180 (38%) |
| **Residence: Health Region (HR)** |  |
| 1st HR of Attica | 299 (68%) |
| 2nd HR of Piraeus & Aegean Islands | 1 (0.2%) |
| 3rd HR of Macedonia | 84 (19%) |
| 4th HR of Macedonia and Thrace | 9 (2.1%) |
| 6th HR of Peloponnese, Ionian islands, Epirus and Western Greece | 17 (3.9%) |
| 7th HR of Crete | 27 (6.2%) |
| Unknown | 38 |
| **Profession** |  |
| Pediatricians | 150 (32%) |
| GPs & Family physicians | 77 (16%) |
| Psychiatrists | 60 (13%) |
| Psychologists | 60 (13%) |
| Nurses | 52 (11%) |
| Child psychiatrists | 40 (8.4%) |
| Social workers<sup>2</sup> | 20 (4.2%) |
| Psych Nurses | 8 (1.7%) |
| Occupational therapists | 5 (1.1%) |
| Rural Doctors | 3 (0.6%) |
| **Type of service(s) they work<sup>2</sup>** |  |
| Primary care | 53 (11%) |
| Community center | 25 (5.3%) |
| Support mobile team | 4 (0.8%) |
| Hospital (outpatient) | 61 (13%) |
| Hospital (inpatient) | 121 (25%) |
| Private practice | 247 (52%) |
| Shelter or NGO | 8 (1.7%) |
| **Working with vulnerable populations<sup>2</sup>** |  |
| Victims of abuse & neglect | 86 (18%) |
| People in refugee camps | 70 (15%) |
| Unaccompanied minors | 45 (9.5%) |
| Roma communities | 68 (14%) |
| Pomak communities | 24 (5.1%) |
| LGBTQI+ communities | 72 (15%) |
| Other specific populations | 338 (71%) |

<sup>1</sup>Mean (SD, Minimum-Maximum); n (%)
<sup>2</sup>Note: percentages do not sum up 100% because educators were allowed to give more than one response. Percentages represent the proportion of the whole sample of educators.

**Table 5.** Mean scores and range of selected instruments measuring mental health symptoms.

| Instrument | Mean scores<br>and score distributions |  |  |  | N and % of children<br>above at risk cut-offs |  |
| --- | --- | --- | --- | --- | --- | --- |
|  | Caregivers |  | Children/adolescents |  | Caregivers | Children/adolescents |
|  | N | Mean (SD, Min-Max) | N | Mean (SD, Min-Max) | n (%) | n (%) |
| CATS-2 | 452 |  | 400 |  |  |  |
| At least one trauma (lifetime) |  |  |  |  | 198 (43,8%) | 137 (34,25%) |
| DSHI |  |  | 414 | 4.9 (25.3, 0.0-365.0) |  |  |
| At least one self-harm episode (six months) |  |  |  |  |  | 56 (13,52%) |
| PSC total emotional or psychosocial problems | 1,356 | 15 (11, 0-68) | 1,201 | 13 (11, 0-70) | 145 (11%) | 100 (8,3%) |
| Attention subscale | 1,356 | 2.54 (2.31, 0.00-10.00) | 1,201 | 2.55 (2.33, 0.00-10.00) | 93 (6,9%) | 82 (6,8%) |
| Internalization subscale | 1,356 | 1.90 (2.07, 0.00-10.00) | 1,201 | 1.83 (2.09, 0.00-10.00) | 164 (12%) | 150 (12%) |
| SDQ total difficulties | 452 | 9 (7, 0-31) | 414 | 8.7 (6.1, 0.0-32.0) | 72 (16%) | 31 (7.5%) |
| Emotional problems | 452 | 2.28 (2.30, 0.00-10.00) | 414 | 2.23 (2.45, 0.00-10.00) | 41 (9,1%) | 19 (4.6%) |
| Conduct problems | 452 | 2.09 (1.91, 0.00-8.00) | 414 | 2.27 (1.67, 0.00-9.00) | 61 (13%) | 20 (4.8%) |
| Hyperactivity | 452 | 3.11 (2.38, 0.00-10.00) | 414 | 2.85 (2.42, 0.00-10.00) | 22 (4,9%) | 18 (4.3%) |
| Peer problems | 452 | 1.97 (1.90, 0.00-9.00) | 414 | 1.37 (1.64, 0.00-9.00) | 59 (13%) | 6 (1.4%) |
| RCADS total score | 452 | 17 (18, 0-112) | 400 | 22 (24, 0-140) |  |  |
| Generalized anxiety | 452 | 2.62 (2.74, 0.00-14.00) | 400 | 3.4 (3.4, 0.0-17.0) | 40 (8,8%) | 62 (16%) |
| Panic symptoms | 452 | 1.84 (3.50, 0.00-23.00) | 400 | 3.0 (4.6, 0.0-27.0) | 16 (3,5%) | 25 (6,2%) |
| Social phobia | 452 | 5.0 (4.4, 0.0-23.0) | 400 | 6.1 (5.5, 0.0-27.0) | 60 (13%) | 79 (20%) |
| Separation anxiety | 452 | 2.8 (3.4, 0.0-18.0) | 400 | 2.8 (3.7, 0.0-21.0) | 95 (21%) | 87 (22%) |
| Obsessive-compulsive symptoms | 452 | 1.86 (2.69, 0.00-15.00) | 400 | 2.4 (3.3, 0.0-18.0) | 65 (14%) | 83 (21%) |
| Depression | 452 | 3.3 (4.4, 0.0-27.0) | 400 | 4.1 (5.4, 0.0-30.0) | 30 (6,6%) | 42 (10%) |
| SNAP total score | 452 | 15 (14, 0-74) |  |  |  |  |
| Inattention | 452 | 6.5 (5.8, 0.0-27.0) |  |  | 55 (12%) |  |
| Hyperactivity | 452 | 4.5 (5.2, 0.00-27.0) |  |  | 32 (7,1%) |  |
| KIDSCREEN quality of life | 452 | 25.4 (4.6, 4.0-37.0) | 400 | 24.6 (5.5, 0.0-37.0) |  |  |

**Abbreviations:** CATS-2 (CHild and Adolescent Trauma Screen-2), DSHI (Deliberate Self Harm Inventory), PSC (Pediatric Symptoms Checklist), RDACS (Revised Children's Anxiety and Depression Scale), SDQ (Strength and Difficulties Questionnaire), SNAP (Swanson, Nolan and Pelham Scale (SNAP-IV).
**Note:** these are unrelated populations, and the ratings by caregivers and by children/adolescents do not correspond to multiple-informant rates for the same individuals.

As for the demographics of participants, the 1.756 caregivers were aged 19 to 73 years-old (mean 41, SD 8), caring for children between 1.1 to 17.9 years (mean 10.1, SD 4.9). Most caregivers were female (54%), and the majority of them (96%) reported to be biological parents of the children/adolescents. The children and adolescents groups summed 1,201 participants aged 8 to 17 years old (median 12.48, SD 2,86), with an even distribution of gender (49% female, 51% male, 0.08% other). The majority of the participants were currently in primary school (44%), followed by gymnasium (28%) and lyceum (24%). Noteworthy, 15% of children/adolescents reported they were told they had a psychological, behavior, or learning problem.

The 404 school teachers aged 25 to 67 years-old (mean 48, SD 10) were predominantly female (69%), worked in public schools (92%) and were evenly distributed across different levels of education (45% taught at primary school, 24% at gymnasium, 25% at lyceum, and 11% at vocational school (EPAL)). They concentrated on the region of Attica (67%). Only 10% of them reported being involved in mental health programs at school. The 475 healthcare professionals were also predominantly female (62%), were aged 28 to 80 years-old (mean 50, SD 9), and were composed of the following specialities: pediatricians (32%), general practitioners and family physicians (16%), psychiatrists (13%), psychologists (13%), nurses (11%), child psychiatrists (8.4%), social workers (4.2%), mental health nursers, occupational therapists (1.1%), and rural doctors (0.6%). They were distributed across different types of services, and private practice concentrated 52% of professionals, followed by inpatient hospital care with 25%. A significant number of them reported working with vulnerable populations (18% with victims of abuse and neglect).

Instruments assessing a range of mental health domains were applied to 1,756 caregivers and 1,201 children (see **Table 5**), providing a screening prevalence for at-risk individuals in this sample. For instance, 8,3% (self-report) to 11% (caregivers-rated) of participants screened positive for emotional or psychosocial problems, 13% are at-risk scores for conduct problems, and 14% presented high levels of obsessive compulsive symptoms. Furthermore, history of lifetime trauma was present for 34% (self-report) to 44% (caregivers-rated) of participants, and 14% reported at least one self-harm episode in the previous six months.

### 3.2) Focus groups

Fourteen focus groups covering research topics were conducted with members from the general community and with professionals from the healthcare and welfare system (see **Table 3** for details of group participants, place, and duration). A summary description of each focus group is now openly available in the Open Science Framework webpage [https://osf.io/crz6h/], and the transcripts can be sent upon request. This material can be used by researchers aiming at investigating specific research questions that guided the development of this dataset, as well as further topics of interest possibly contained within the richness of focus groups. Initial qualitative analysis is shown in **Table 6**, which outlines the key findings for overarching groups (refer to **Supplementary Table 9** for data extracts supporting each finding).

**Table 6.** Key initial findings from focus groups.

| Children and adolescents | Minority groups (LGBTQIA+, Pomak, Roma, and refugee adolescents) |
| --- | --- |
| • Children and adolescents believe mental health problems appear through actions and appearance, including rudeness, avoiding activities, or crying. However, some perceive such struggles may not be apparent to others or themselves.<br>• They note the role of external factors on mental health, but acknowledge individual factors as well. They recognize vignettes of depression and ADHD, and even mention conditions like dyslexia.<br>• Stigmatized views are more disseminated among children and adolescents, as discrediting disorders, believing people with depression should just feel better, and belittling symptoms.<br>• Peer stigma is a barrier to seeking help, as many would not want their peers to know they are seeing a professional. They believe seeing others using mental health services could help normalize care.<br>• Children and adolescents identify family and specialists as sources for help, and assume some problems need to be addressed by specialists.<br>• Sometimes there is a disbelief that help could make them feel better or doubts they would be able to recognize help is needed. | • LGBTQIA+ adolescents conceptualize that mental health problems may be experienced privately and not shown out.<br>• When conceptualizing wellness, Pomak, refugees, and LGBTQIA+ adolescents referred to the importance of freedom and feeling free.<br>• Some minority groups are especially vulnerable to stigmatized views. Refugee minors conceptualize mental health issues as “craziness” to be dealt with by deprivation in psychiatric hospitals. Pomak and Roma adolescents believe people with mental health problems should “distract” themselves and not seek professional assistance.<br>• Roma adolescents are less likely to seek assistance because they would not want to seek help outside of their community. |
| Caregivers | Teachers and healthcare professionals |
| • Caregivers are aware of the importance of children and adolescents’ mental health, being willing to discuss and to engage in learning about it.<br>• Caregivers are confident that they can identify mental health problems in their kids through signs such as behavior, uncontrolled emotions, shifts in routine, and social isolation.<br>• They are aware of the importance of the environment in mental health. Signs of mental health struggles are attributed | • There is a positive attitude towards mental health among all professional categories, who feel responsible for caring for children and adolescents.<br>• They understand that environmental factors are determinants of mental health, including violence at home, parental neglect, or school problems. They are also aware of specific struggles faced by minorities, including race, sexuality, and disability. |
| to stressors such as suffering from love, isolation, fights, social media, or challenges at school.<br>• Caregivers did not perform well in identifying mental health conditions when shown vignettes. Typical signs of ADHD were not recognized, having this manifestation attributed to issues at home or to the overuse of technology.<br>• There is a willingness to accept psychotherapy, but parents are reluctant towards prescription drugs.<br>• For caregivers, the initial step to addressing mental health issues is to try and solve it themselves or with advice from other parents, only reaching out for specialists if the issue persists.<br>• Caregivers recognize that taboo may be a barrier to seek for help, but none recognize it as a barrier for themselves.<br>• Several caregivers spoke of their personal lack of understanding and struggles opening up about mental health. | • Healthcare providers see both parents and children struggling to understand the different facets of mental health, with limited awareness of the available resources. They believe more information for the general public is crucial.<br>• Health professionals see stigma as a problem in Greece, especially among adults. The discomfort when talking about mental health would hold people from seeking help. This is especially pronounced in remote areas, with rural inhabitants even looking for assistance in urban areas as they worry over being perceived as weak.<br>• Teachers note that peer and school community acceptance or stigmatization is a challenge, and point out that students fear friends will find out they are seeking assistance.<br>• Teachers and health professionals stress out that one of the biggest challenges is overcoming parental refusal that care is needed. Health providers attribute it to not knowing treatment options and fearing what they may entail.<br>• Healthcare professionals believe it is necessary to improve relationships between caregivers and children/adolescents so that they feel more comfortable expressing their struggles. |

**Abbreviations:** ADHD (Attention Deficit/Hyperactivity Disorder).

Among children and adolescents, a prevailing finding was the presence of stigma regarding mental health. This could be expressed in the form of internalized stigma, as in conceptions that shame or belittle mental health problems, but also played out in social dynamics, and peer shaming proved a significant barrier for reaching out for help. Whilst the findings suggest children/adolescents present a certain degree of literacy for recognizing common mental health conditions, they also struggle in self-recognition of symptoms and do not know available treatments and their possible benefits. A common view held by children and adolescents was that nothing could make them feel better in case of facing mental health symptoms, summing another obstacle for seeking assistance. Moreover, the findings from minority groups revealed special needs to be considered in these populations. These groups were especially vulnerable to stigmatized views of mental health, including extreme ones such as mental health problems as craziness to be dealt in reclusion hospitals. Also, specific struggles were apparent when they conceptualized wellness, such as the need to feel free in order to feel well, which was elaborated by LGBTQIA+, Pomak, and refugee participants. These groups also face special barriers for mental health care access, as Roma adolescents revealed they would be reluctant in reaching out for help outside the community as a result of lack of trust.

As for the caregivers, positive attitudes were found regarding mental health problems, with awareness of its importance and openness for discussing the subject. Nevertheless, this was counteracted by a limitation on their literacy on mental health: caregivers overemphasized external issues as the only source of mental health struggles, and could not recognize typical conditions such as Attention Deficit/Hyperactivity Disorder (ADHD). This ultimately led to stigmatized attitudes on mental health, as a reluctance towards medication treatment, an overreliance in their capacity of addressing possible issues by themselves, and a closeness in speaking out of their own mental health difficulties.

The findings from teachers and healthcare professionals were further informative of the struggles faced by children, adolescents, and caregivers. From where they stand, these professionals perceive adults and adolescents alike struggling to understand their mental health demands and which resources are available. They endorse that stigma and literacy are crucial issues, further revealing a challenge on overcoming parental resistance that there might be a problem, and a school climate that leads students to fear peer stigma. The taboo is perceived to be more pronounced among the adult population (especially in rural areas), which struggle to deal with their own mental health and also resist in addressing the mental health of their children, frequently fearing what treatment options may entail or considering mental health issues as weakness. This way, children/adolescents do not encounter a welcoming environment for reporting their issues either at home or at school.

## 4. Discussion

We report the development of an open science repository through a mixed-method, community-based design to assess the current state and needs of child and adolescent mental health in Greece according to the perspective of multiple informants. The repository can be freely navigated at [https://osf.io/crz6h/], and encompassess the datasets of the survey with 1,756 caregivers, 1,201 children/adolescents, 404 teachers, and 475 healthcare professionals, covering measures of mental health conditions, mental health needs, literacy and stigma, service use and access, professional practices, professional training background, and training needs and preferences. It also contains material from 14 focus groups presenting in-depth explorations on such topics. We expect the repository to have many applications for upcoming research, as it can be approached by scholars and policymakers to investigate diverse questions to understand needs and priorities for child and adolescent mental health. Within the CAMHI, this dataset will ground several evidence-based projects, including a tailored training program for health professionals and teachers across Greece, the implementation of a nationwide network referral and supervision systems for mental health assistance, and the development of online resources for the community aimed at mental health promotion.

This work draws on principles of dissemination and implementation research, as it underpins the construction of an open science repository aimed at providing evidence-based information to improve the mental health care of a given real-world population (Shelton et al. 2020). This way, the research was oriented to practice from its conception, establishing strong links between these poles that are often gapped. For instance, the results from the questionnaires assessing the background training, skills, and training needs and perspectives from professionals have direct application in tailoring a training program that may effectively improve mental health assistance. This highlights the importance of conducting contextualized, engaged research, as it produces results that have immediate applicability, potentializing impacts and optimizing resources. It also denotes that policymaking and intervention programs are feasible to be conducted within validated research paradigms, making a further point to evidence-based initiatives. This work also abides to the principles of open science, making all resources freely available for the general and professional communities. Beyond increasing the outreach and real-world impact, this approach also enhances the credibility and reproducibility of research, as data can be thoroughly accessed by peers (Tennant et al. 2016).

This study has many strengths in its design. By applying a mixed-method design, it may unveil complementary aspects of the phenomenon that are not always captured using a single method (Creswell and Clark 2017). Furthermore, both quantitative and qualitative arms are in accordance with established standards of research and were reported according to validated guidelines in their fields, namely the COREQ and the STROBE (Tong, Sainsbury, and Craig 2007; von Elm et al. 2014). Another strength is the diversity of viewpoints obtained by the participation of multiple stakeholders, which can enrich findings. This is especially relevant when conducting focus groups with vulnerable groups, guaranteeing a voice for underrepresented minorities (namely Roma, Pomak, refugee, and LGBTQIA+ adolescents). In this same vein, the work also benefits from a wideness of scope, with multiple yet pertinent topics being inquired and having the potential to compose a comprehensive map of the scenario under analysis.

We also face limitations. Apart from the group of children/adolescents recruited by random telephone calls, our samples were non-probabilistic as they were based on third-party panels matched for demographic variables according to the census distribution. Therefore, they are not intended at estimating the national prevalence of conditions. The recruitment of teachers does not represent distant areas, as it was done in major cities in a face-to-face fashion. The inquiry with caregivers used a third-party online respondent panel that faced a maximum number of participants, restricting greater sample sizes. Schoolteachers, healthcare professionals, and one of the groups of children and adolescents were surveyed using a telephone closed-end interview. This procedure increases completion of questionnaires, but also introduces some inherent biases that are not expected in self-applied instruments. Moreover, we had low response rates across some groups, which were especially for children and adolescents recruited via random landline phone calls (4,74%). This might be attributed to our recruitment strategy, as the need for caregivers’ consent implied further procedures that diminished response, and random phone line calls are expected to return low completion rates. We also lack the characteristics of people who refused to participate, precluding adequate comparison with participants. Concerning the qualitative part, focus groups are a recommended methodology for raising perceptions of shared experiences, as discussions combine multiple perspectives that would hardly be reached by individual interviews (Casey 2009). Nevertheless, participants who respond to invitations may have inclinations to be risk takers or be more assertive than non-participants. It is also common that some participants speak more forcefully during sessions, increasing the weight of their opinions in relation to others. Individuals with mental health issues may also be less inclined to participate in focus groups, and so their views might be underrepresented.

This open repository can now be freely navigated at [https://osf.io/crz6h/], containing valuable information from multiple stakeholders on key topics for child and adolescent mental health in Greece. We expect it to be used by the professional and general community to embase upcoming research and policy making. This initiative may encourage similar ones aiming at generating positive impact for mental health by designing and disseminating research, providing a methodological strategy that can ground related projects in other countries.

## Data Availability

Most data that support the findings of this study are openly available in Open Science Framework at http://doi.org/10.17605/OSF.IO/CRZ6H. Raw data are currently available from the corresponding author [GAS] on request, and will be made freely available after completing one year of data collection.

## Acknowledgments

The authors would like to thank the Stavros Niarchos Foundation (SNF) for funding the SNF-CMI Child and Adolescent Mental Health Initiative and SNF’s Co-President Andreas C. Dracopoulos for his leadership in creating, launching, and supporting the project. We would also like to thank Ms. Elianna Konialis, Ms. Dimitra Moustaka and Mr. Panos Papoulias for their critical role in multiple steps of the conceptualization and implementation of the SNF-CMI Child and Adolescent Mental Health Initiative. We also thank Samanta Duarte for designing the graphical representations included in this paper.

