## Supplementary for "Understanding priorities and needs for child and adolescent mental health in Greece from multiple informants: an open resource"


| | |
|---|---|
| Supplementary Table 1. Strengthening the reporting of observational studies in epidemiology (STROBE) checklist | 1 |
| Supplementary Table 2. Consolidated criteria for reporting qualitative studies (COREQ) checklist | 4 |
| Supplementary Table 3. Description of selected instruments for the survey | 6 |
| Supplementary Table 4. Five-stage procedure to cross-cultural adaptation of instruments | 9 |
| Supplementary Table 5 - Quotas for the survey | 10 |
| Supplementary Table 6. Detailed recruitment procedure for focus groups | 12 |
| Supplementary Table 7. Mental health vignettes depicting prevalent conditions for each age group | 17 |
| Supplementary Table 8. Structure of the guiding manuals for focus groups | 18 |
| Supplementary Table 9. Extracts supporting focus groups’ findings | 20 |

**Supplementary Table 1. Strengthening the reporting of observational studies in epidemiology (STROBE) checklist**

| | | Recommendation | Page |
|---|---|---|---|
| **Title and abstract** | 1 | (*a*) Indicate the study's design with a commonly used term in the title or the abstract | #3 |
| | | (*b*) Provide in the abstract an informative and balanced summary of what was done and what was found | #3 |
| Introduction | | | |
| Background /rationale | 2 | Explain the scientific background and rationale for the investigation being reported | #4 |
| Objectives | 3 | State specific objectives, including any prespecified hypotheses | #5 |
| Methods | | | |
| Study design | 4 | Present key elements of study design early in the paper | #5 |
| Setting | 5 | Describe the setting, locations, and relevant dates, including periods of recruitment, exposure, follow-up, and data collection | #5 - #8 |
| Participants | 6 | (*a*) Give the eligibility criteria, and the sources and methods of selection of participants | #6 - #8<br>Figure 1<br>Figure 2 |
| Variables | 7 | Clearly define all outcomes, exposures, predictors, potential confounders, and effect modifiers. Give diagnostic criteria, if applicable | #6 - #8 |
| Data sources/ measurement | 8* | For each variable of interest, give sources of data and details of methods of assessment (measurement). Describe comparability of assessment methods if there is more than one group | #6<br>Table 1<br>Table 2<br>Supplementary Table 3<br>Supplementary Table 4 |
| Bias | 9 | Describe any efforts to address potential sources of bias | #6 - #8 |
| Study size | 10 | Explain how the study size was arrived at | #6 - #8 |
| Quantitative variables | 11 | Explain how quantitative variables were handled in the analyses. If applicable, describe which groupings were chosen and why | #8 |

| | | | |
|---|---|---|---|
| Statistical methods | 12 | (*a*) Describe all statistical methods, including those used to control for confounding | #8 |
| | | (*b*) Describe any methods used to examine subgroups and interactions | #8 |
| | | (*c*) Explain how missing data were addressed | Not applicable |
| | | (*d*) If applicable, describe analytical methods taking account of sampling strategy | Not applicable |
| | | (*e*) Describe any sensitivity analyses | Not applicable |
| Results | | | |
| Participants | 13* | (a) Report numbers of individuals at each stage of study—eg numbers potentially eligible, examined for eligibility, confirmed eligible, included in the study, completing follow-up, and analysed | #6-8 |
| | | (b) Give reasons for non-participation at each stage | Not applicable |
| | | (c) Consider use of a flow diagram | Not applicable |
| Descriptive data | 14* | (a) Give characteristics of study participants (eg demographic, clinical, social) and information on exposures and potential confounders | Table 4 |
| | | (b) Indicate number of participants with missing data for each variable of interest | Not applicable |
| Outcome data | 15* | Report numbers of outcome events or summary measures | Not applicable |
| Main results | 16 | (*a*) Give unadjusted estimates and, if applicable, confounder-adjusted estimates and their precision (eg, 95% confidence interval). Make clear which confounders were adjusted for and why they were included | Not applicable |
| | | (*b*) Report category boundaries when continuous variables were categorized | Not applicable |
| | | (*c*) If relevant, consider translating estimates of relative risk into absolute risk for a meaningful time period | Not applicable |
| Other analyses | 17 | Report other analyses done—eg analyses of subgroups and interactions, and sensitivity analyses | Not applicable |
| Discussion | | | |
| Key results | 18 | Summarise key results with reference to study objectives | Not applicable |

| | | | |
|---|---|---|---|
| Limitations | 19 | Discuss limitations of the study, taking into account sources of potential bias or imprecision. Discuss both direction and magnitude of any potential bias | #15 |
| Interpretati on | 20 | Give a cautious overall interpretation of results considering objectives, limitations, multiplicity of analyses, results from similar studies, and other relevant evidence | Not applicable |
| Generalisabi lity | 21 | Discuss the generalisability (external validity) of the study results | Not applicable |
| Other information | | | |
| Funding | 22 | Give the source of funding and the role of the funders for the present study and, if applicable, for the original study on which the present article is based | #2 |

**Supplementary Table 2. Consolidated criteria for reporting qualitative studies (COREQ) checklist**

| No. Item | Guide questions/description | Reported on Page # |
|---|---|---|
| **Domain 1: Research team and reflexivity** | | |
| *Personal Characteristics* | | |
| 1. Interviewer - facilitator | Which author/s conducted the interview or focus group? | Supplementary Table 6 |
| 2. Credentials | What were the researcher's credentials? (e.g., PhD, MD) | Supplementary Table 6 |
| 3. Occupation | What was their occupation at the time of the study? | Supplementary Table 6 |
| 4. Gender | Was the researcher male or female? | Supplementary Table 6 |
| 5. Experience and training | What experience or training did the researcher have? | #9<br>Supplementary Table 6 |
| *Relationship with participants* | | |
| 6. Relationship established | Was a relationship established prior to study commencement? | #9 |
| 7. Participant knowledge of the interviewer | What did the participants know about the researcher? (e.g., personal goals, reasons for doing the research, etc.) | #9 |
| 8. Interviewer characteristics | What characteristics were reported about the interviewer/facilitator? (e.g., bias, assumptions, reasons and interests in the research topic) | #9 |
| **Domain 2: study design** | | |
| *Theoretical framework* | | |
| 9. Methodological orientation and Theory | What methodological orientation was stated to underpin the study? (e.g., grounded theory, discourse analysis, ethnography, phenomenology, content analysis) | Not applicable |
| *Participant selection* | | |
| 10. Sampling | How were participants selected? (e.g., purposive, convenience, consecutive, snowball) | #9<br>Supplementary Table 6 |
| 11. Method of approach | How were participants approached? (e.g., face-to-face, telephone, mail, email) | #9<br>Supplementary Table 6 |
| 12. Sample size | How many participants were in the study? | #8<br>Supplementary Table 6 |
| 13. Non-participation | How many people refused to participate or dropped out? Reasons? | Supplementary Table 6 |
| *Setting* | | |
| 14. Setting of data collection | Where was the data collected? (e.g., home, clinic, workplace) | #8 |
| 15. Presence of non-participants | Was anyone else present besides the participants and researchers? | #9 |

| | | |
|---|---|---|
| 16. Description of sample | What are the important characteristics of the sample? (e.g., demographic data, date) | #8<br>Table 3<br>Supplementary Table 6 |
| *Data collection* | | |
| 17. Interview guide | Were questions, prompts, guides provided by the authors? Was it pilot tested? | #10<br>Supplementary Table 8 |
| 18. Repeat interviews | Were repeat interviews carried out? If yes, how many? | Not applicable |
| 19. Audio/visual recording | Did the research use audio or visual recording to collect the data? | #9 |
| 20. Field notes | Were field notes made during and/or after the inter view or focus group? | #9 |
| 21. Duration | What was the duration of the interviews or focus group? | #8 |
| 22. Data saturation | Was data saturation discussed? | Not applicable |
| 23. Transcripts returned | Were transcripts returned to participants for comment and/or correction? | #9 |

**Supplementary Table 3. Description of selected instruments for the survey**

| Instrument | Description |
|---|---|
| *Experience of Service Questionnaire (ESQ)* | A 12-item instrument measuring service satisfaction in child and Adolescent Mental Health Service. |
| *The Mental Health Literacy Scale (MHLS)* | A 35-item questionnaire that measures six attributes: (1) ability to recognize mental disorders, (2) knowledge of risk factors and causes for mental disorders, (3) knowledge of self-treatment, (4) knowledge of professional help available, (5) knowledge of where to seek information, and (6) attitudes that promote recognition or appropriate help-seeking behavior. Items are measured on Likert scale of 4 to 5 points. |
| *The Reported and Intended Behavior Scale (RIBS)* | An 8-item scale evaluating respondent's previous social contact experience and intentions towards people with mental health problems. Items are measured on a 5-point Likert scale. |
| *The Barriers to Access to Care Evaluation (BACE)* | A 30-item scale assessing and identifying key barriers to care experienced by people who currently use, or have used in the past, mental health services. Items are measured on a 4-point Likert scale. Instrument modified with the permission of King's College London. |
| *Pediatric Symptoms Checklist (PSC)* | A 35-item measure used to identify youth's emotional and behavioral adjustment and is meant to provide an assessment of psychosocial functioning. Respondents rate on a 3-point Likert scale how often they experience the emotional or behavioral problems being described. The instrument yields a total score that indicates psychological difficulties, as well as three scores on the subscales of attention, anxiety/depression (internalizing problems), and conduct/disruptive behavior (externalizing problems). |
| *Strength and Difficulties Questionnaire (SDQ)* | A 25-item emotional and behavioral questionnaire designed to capture the perspective of children, young people, parents, and teachers about the children and adolescents' emotional symptoms, conduct problems, hyperactivity/inattention, peer relationship problems, and prosocial behavior. It can be completed by children aged 11 to 17 years old and by parents of children and adolescents aged 2 to 17 years old. |
| *Modified Checklist for Autism in Toddlers, Revised (M-CHAT)* | A 20-item tool for early detection of signs of Autism Spectrum Disorder (ASD) among toddlers assessing overall neurodevelopment, social interaction, and repetitive behaviors, among other typical ASD symptoms. |
| *Childhood Autism Spectrum Test (CAST)* | A 37-item parental report questionnaire developed to detect subtler manifestations of Autism Spectrum Conditions (ASC), including Asperger Syndrome (AS) in primary school children. It measures difficulties and preferences in social and communication skills covering (1) initiation and maintenance of conversation and specific language difficulties (2) social interaction with adults and peers, including eye contact, (3) choice of play |

| | |
|---|---|
| | activities, (4) presence of rigid or repetitive behaviors, (5) choice of interests and sharing interests with others. |
| *Parent Behavior Inventory (PBI)* | A 20-item scale used to measure parenting behavior towards preschool-age and young school-age children. The instrument has two independent scales, Supportive/Engaged and Hostile/Coercive. |
| *Revised Children's Anxiety and Depression Scale (RCADS-47)* | A 47-item scale that measures levels of anxiety and low mood. It includes 6 subscales: separation anxiety disorder, social phobia, generalized anxiety disorder, panic disorder, obsessive compulsive disorder, and low mood (major depressive disorder). It also yields a Total Anxiety Scale (sum of the 5 anxiety subscales) and a Total Internalizing Scale (sum of all 6 subscales). Items assess the frequency of symptoms and are rated on a 4-point Likert scale. |
| *KIDSCREEN-10* | A well-being and health-related quality of life (HRQoL) measure consisting of 10 items answered on a 5-point Likert scale. The items explore the level of the child/adolescent's physical activity, energy, and fitness, depressive moods and emotions, opportunities to enjoy leisure and social activities, quality of interaction with parents and peers, and capacity and satisfaction with school performance. Higher values indicate better health-related quality of life. |
| *Child and Adolescent Trauma Screen-2* (CATS-2) | A short questionnaire aimed to screen for exposure to potentially traumatic events and Posttraumatic Stress Disorder (PTSD) symptoms. The instrument is based on the DSM-5 criteria for PTSD and the items encompass symptoms of intrusions, avoidance, negative alterations in cognitions and mood and hyperarousal. Traumatic events are elicited using a 15-item checklist, followed by an item asking which traumatic experience was more disturbing. If there is at least one disturbing traumatic exposure, PTSD symptoms are measured by 20 items rated on a 4-point Likert scale. |
| *ABCD Parent Screen Use questions* | Four questions were included to assess children's average daily time spent on electronic devices, texting, video chatting, and on social media over the period of a week. |
| *CRAFFT Screening Tool For Substance Use Disorders* (CRAFFT 2.1+N) | A health screening tool designed to identify substance use, substance-related riding/driving risk, and substance use disorder among adolescents. Information yielded can serve as the basis for early intervention. It is composed of 21 self-reported binary items about substance use and associated problems and impairment. |
| *Deliberate Self-Harm Inventory – 9 items (DHSI-9)* | An 9-item measure of various aspects of NSSI originally developed for use with adults that is adapted for use with adolescents. It assesses the presence and frequency of the most common forms of NSSI in adolescents, including cutting, burning, severe scratching, self-biting, carving, sticking sharp objects into the skin, self-punching, and head banging (all to the extent that scarring, bleeding, and/or bruising occurred). |
| *ABCD Screen Use questions* | Eleven questions were included to assess children's average daily time spent watching movies, videos, or TV, playing video games, texting, visiting social |

| | |
|---|---|
| | media, video chatting, browsing the internet, and talking to friends over the period of a week. |
| *The Multitheoretical List of Interventions-30 Items* (MULTI-30) | The MULTI-30 assesses how frequently professionals used 30 psychotherapy techniques on the last six months through a 5-point Likert scale ranging from *"never/almost never"* to "*always/almost always".* The instrument is divided into eight subscales representing (a) common psychotherapy factors and the therapeutic approaches of (b) psychodynamic therapy (c) process-experiential psychotherapy (d) interpersonal psychotherapy (e) person-centered psychotherapy (f) behavioral therapy (g) cognitive therapy, and (h) dialectical behavioral therapy). |
| *Swanson, Nolan and Pelham Scale (SNAP-IV)* | A 26-item instrument screening symptoms of hyperactivity, inattention, impulsivity, and oppositional behavior. Questions are answered using a Likert-scale, and risk scores are calculated by summing the punctuation in each symptom cluster, with higher scores pointing to higher levels of symptoms. |

**Supplementary Table 4. Five-stage procedure to cross-cultural adaptation of instruments**

| Stage | Procedure |
|---|---|
| 1 | Two different translators with expertise in the field will independently translate the instruments into the Greek language |
| 2 | Discrepancies between the two versions will be resolved to create a synthesized Greek version of the instrument |
| 3 | Two different translators naïve to the measured outcomes will back-translate the instrument to English |
| 4 | An expert committee will evaluate semantic, idiomatic, experiential, and conceptual equivalence across all the versions of the instrument and general a prefinal version |
| 5 | The pre final version is tested on the target population |

**Note**: refer to the full guideline for a complete description and rationale of procedure (Beaton et al. 2000).

**Supplementary Table 5 - Quotas for the survey**

| Caregivers of children aged 18 to 30 months | | | Caregivers of children aged 3 to 17 years-old | | | Children/adolescents (Group 2, recruited via random phone calls) | | |
|---|---|---|---|---|---|---|---|---|
| Region | Target quota | Achieved quota | Region | Target quota | Achieved quota | Region | Target quota | Achieved quota |
| Attiki | 46,60% | 47% | Attiki | 44,70% | 45,1% | Attiki | 36,00% | 38% |
| Central - East Macedonia & Thrace / Thessaloniki | 24,30% | 24,5% | Central - East Macedonia & Thrace / Thessaloniki | 24,70% | 25,6% | Central - East Macedonia & Thrace / Thessaloniki | 23,00% | 23% |
| West Macedonia / Epirus | 5,00% | 5% | West Macedonia / Epirus | 5,40% | 5,7% | West Macedonia / Epirus | 6,00% | 5% |
| Thessaly / Central Greece | 6,20% | 6% | Thessaly / Central Greece | 8,30% | 8,3% | Thessaly / Central Greece | 11,00% | 11% |
| West Greece / Peloponnese / Ionian Islands | 7,50% | 7,5% | West Greece / Peloponnese / Ionian Islands | 8,10% | 7,4% | West Greece / Peloponnese / Ionian Islands | 13% | 12% |
| Aegean islands / Crete | 10,40% | 10% | Aegean islands / Crete | 8,80% | 7,8% | Aegean islands / Crete | 11% | 11% |
| Child's gender | | | Child's gender | | | Gender | | |
| Male | 50,00% | 50% | Male | 52,00% | 52,9% | Male | 52% | 49% |
| Female | 50,00% | 50% | Female | 48,00% | 47% | Female | 48% | 51% |
| Other | NA | NA | Other | NA | 0,1% | Other | NA | NA |
| Income | | | Income | | | Age | | |
| Low income* | 58,00% | 60% | Low income* | 49,00% | 51,9% | 8 - 12 years | 49% | 54% |
| Medium/High income* | 28,00% | 36% | Medium/High income* | 37,00% | 43% | 13 - 17 years | 51% | 46% |
| Prefer not to answer | 14% | 4% | Prefer not to answer | 14% | 5,2% | | | |

| Teachers and healthcare professionals | | | | | | | | | | |
|---|---|---|---|---|---|---|---|---|---|---|
| | **Attiki** | | **Evros** | | **Heraklion** | | **Thessaloniki** | | **Ioannina** | |
| | Target | Achieved | Target | Achieved | Target | Achieved | Target | Achieved | Target | Achieved |
| **Teachers** | 67,2% | 67,6% | 2,6% | 2,6% | 6,8% | 6,7% | 20,3% | 20% | 3,2% | 3,2% |
| **Child** | 69% | 70% | 2% | 3% | 5% | 5% | 20% | 20% | 3% | 3% |

| | | | | | | | | | | |
|---|---|---|---|---|---|---|---|---|---|---|
| **psychiatrists** | | | | | | | | | | |
| **GPs & Family physicians** | 66% | 66% | 3% | 3% | 7% | 8% | 20% | 20% | 4% | 4% |
| **Nurses** | 61% | 62% | 3% | 2% | 5% | 6% | 24% | 23% | 7% | 8% |
| **Occupational therapists** | 67% | 63% | 1% | 0% | 5% | 13% | 21% | 25% | 5% | 0% |
| **Pediatricians** | 68% | 80% | 2% | 0% | 7% | 0% | 20% | 20% | 3% | 0% |
| **Psychiatrists** | 72% | 68% | 1% | 2% | 3% | 7% | 21% | 20% | 3% | 3% |
| **Psychologists** | 69% | 72% | 1% | 2% | 5% | 3% | 24% | 20% | 1% | 3% |
| **Social workers** | 60% | 68% | 5% | 2% | 14% | 5% | 18% | 23% | 4% | 2% |
| **Mental health nurses** | 62% | 60% | 3% | 5% | 5% | 10% | 24% | 20% | 5% | 5% |

**Note**: Quotas were established based on the census data (for caregivers, children and adolescents, and teachers) and IQVIA database profiling data (for healthcare professionals) (Hellenic Statistical Authority 2012; IQVIA n.d.). However, socio economics quotas consist of an approximation that considers a range of factors and do not represent official data.

**Supplementary Table 6. Detailed recruitment procedure for focus groups**

| Focus Group No. | Moderator | Approach | Steps | Candidates |
|---|---|---|---|---|
| **FG1. Parents of 15-year-old adolescents (general population)** | ⟶ Psychologist and Researcher, MSc, PhD -> Male<br>⟶ Deputy Manager of the General Hospital of Patras<br>⟶ Project Manager of the Association of Regional Development and Mental Health (EPAPSY) in collaboration with UNHCR | ⟶ Face-to-face contact<br>⟶ Searched online databases for schools, parents, and guardians' associations<br>⟶ Communicated with Directors and Secretaries of public and private schools<br>⟶ Snowball sampling | ⟶ Called candidates to provide further information about the program<br>⟶ sent recruitment questionnaire for completion<br>⟶ Selected candidates who met the criteria<br>⟶ Sent consent forms and received them once signed<br>⟶ Sent candidates the meeting link | Total calls: 37<br>Total emails: 69<br>Reasons for exclusion:<br>⟶ No answer<br>⟶ Children's age did not match the criteria<br>⟶ Unwillingness to participate (vacation period, no interest)<br>⟶ Sample's gender variety |
| **FG2. Parents of 8-year-old children (general population)** | | ⟶ Face-to-face contact<br>⟶ Searched online databases for schools, parents, and guardians' associations<br>⟶ Communicated with Directors and Secretaries of public and private schools<br>⟶ Snowball sampling | Same as in previous focus group | Total calls: 30<br>Total emails: 67<br>Reasons for exclusion:<br>⟶ No answer<br>⟶ Children's age did not match the criteria<br>⟶ Unwillingness to participate (vacation period, no interest)<br>⟶ Sample's gender variety |
| **FG3. Teachers** | | ⟶ Communicated with public, private, and special education schools<br>⟶ Reached out to third parties (e.g., Professionals, Ministry of Education, Unions and Associations)<br>⟶ Snowball sampling | Same as in previous focus group | Total calls: 38<br>Total emails: 67<br>Reasons for exclusion:<br>⟶ No answer<br>⟶ Sample's location variety<br>⟶ Unwillingness to participate (vacation period, no interest) |
| **FG4. Nurses and nurses' assistants** | | ⟶ Communicated with private clinics, | Same as in previous focus group | Total calls: 30<br>Total emails: 61 |

| | | | | |
|---|---|---|---|---|
| | | public hospitals, Public health centers, and Hellenic Nurses' Association<br>⟶ Snowball sampling | | Reasons for exclusion:<br>⟶ Sample's location variety<br>⟶ Unwillingness to participate (vacation period, heavy workload due to COVID-19, rolling working hours) |
| **FG5. Speech and Occupational Therapists** | | ⟶ Communicated with private clinics, the Panhellenic Association of Logopedists, and Speech Therapy Centers<br>⟶ Snowball sampling | Same as in previous focus group | Total calls: 25<br>Total emails: 35<br>Reasons for exclusion:<br>⟶ Sample's location variety<br>⟶ Unwillingness to participate (vacation period, rolling working hours) |
| **FG6. Psychologists and Social Workers** | | ⟶ Communicated with private clinics, NGOs, Child and Adolescent Psychiatrists Association, special education schools, and health centers | Same as in previous focus group | Total calls: 25<br>Total emails: 29<br>Reasons for exclusion:<br>⟶ Sample's location variety<br>⟶ Not specializing in children<br>⟶ Unwillingness to participate (vacation period, rolling working hours) |
| **FG7. Child Psychiatrists, Pediatricians and Family Doctors** | | ⟶ Communicated with private clinics, public hospitals, health centers<br>⟶ Searched online databases | Same as in previous focus group | Total calls: 85<br>Total emails: 105<br>Reasons for exclusion:<br>⟶ Family doctors with no specialization in children or experience<br>⟶ Unwillingness to participate (vacation period, rolling working hours) |
| **FG8. NGO Leaders** | | ⟶ Communicated | Same as in previous | Total calls: 40 |

| | | | | |
|---|---|---|---|---|
| | | with NGO Organizations and third parties<br>⟶ Snowball sampling | focus group | Total emails: 79<br>Reasons for exclusion:<br>⟶ No specialization in children<br>⟶ Unwillingness to participate (vacation period, personal reasons) |
| **FG9. 15-year-old Adolescents (general population)** | ⟶ Psychologist and Researcher, MSc<br>-> Female<br>⟶ Affiliate Institution: National and Kapodistrian University of Athens | ⟶ Face-to-face contact<br>⟶ Searched online databases for schools, parents, and guardians' associations<br>⟶ Communicated with Directors and Secretaries of public and private schools<br>⟶ Snowball sampling | Same as in previous focus group | Total calls: 35<br>Total emails: 72<br>Reasons for exclusion:<br>⟶ Sample's location variety<br>⟶ Unwillingness to participate (limited free time, no interest) |
| **FG10. Roma Adolescents** | | ⟶ Contacted Cultural Associations to present the program and discuss the possibility of access to this particular population<br>⟶ Recruited a cultural mediator from the Association of Greek Roma Cultural Mediators | ⟶ The cultural mediator was in charge of the recruitment of members of a Roma community, based in Athens<br>⟶ The cultural mediator collected the consent forms and the recruitment grid, and escorted children to and from the venue | N/A |
| **FG11. Refugee unaccompanied minors** | | ⟶ Addressed an official request for support to the Special Secretariat for the protection of unaccompanied minor refugees, at the Ministry of Migration and Asylum<br>⟶ Filed request to | ⟶ Mental health professionals of the 2 participating shelters conducted the recruitment of children based on the given criteria (age 13-17, gender representativeness, Farsi speakers, > 2-year-stay in | N/A |

| | | | | |
|---|---|---|---|---|
| | | the Public Prosecutor for minors (Athen’s Court of Firts) for the approval of the Investigaton<br>⟶ Reached out to shelters<br>⟶ Two participating shelters filed a request for participation in the Prosecutor’s office by Hospitality structure | Greece), collected the signed Informed Consent Forms (adapted for this population)<br>⟶ Shelters provided escorts for the supervision and safe transport of children to and from the venue | |
| **FG12. 8-year-old Children (general population)** | | ⟶ Face-to-face contact<br>⟶ Searched online databases for schools, parents, and guardians’ associations<br>⟶ Communicated with Directors and Secretaries of public and private schools<br>⟶ Snowball sampling | ⟶ Called candidates to provide further information about the program<br>⟶ sent recruitment questionnaire for completion<br>⟶ Selected candidates who met the criteria<br>⟶ Sent consent forms and received them once signed<br>⟶ Sent candidates the meeting link | Total calls: 25<br>Total emails: 52<br>Reasons for exclusion:<br>⟶ Children’s age did not match the criteria<br>⟶ Unwillingness to participate (no interest) |
| **FG13. Pomak Adolescents** | Consulting Psychology MSc, PhD Candidate Female<br>⟶ Affiliate Institution: National and Kapodistrian University of Athens | ⟶ Contacted Cultural Associations to present the program and discuss the possibility of access to this particular population<br>⟶ Recruited a cultural mediator | ⟶ Recruitment was similar to FG10.<br>⟶ Two cultural mediators were in charge of the recruitment of children located in a Pomak village outside the city of Xanthi<br>⟶ The cultural mediators collected the consent forms and the recruitment grid, and escorted children and their parents to and from the venue | |

| | | | | |
|---|---|---|---|---|
| **FG14. LGBTQIA+ Adolescents** | | ⟶ Face-to-face contact with community representatives<br>⟶ Communicated with LGBTQIA+ Organizations<br>⟶ Contacted Psychologists, Occupational Therapists, Social Workers, Speech and Language Therapists from previous Groups<br>⟶ Realised a closed social media campaign through private channels and groups<br>⟶ Snowball sampling | ⟶ Recruitment started by contacting third party Organizations of the LGBTQIA+ community (8 NGO's)<br>⟶ After the initial low interest of the Organizations, face-to-face meetings were conducted with NGO's Representatives, parents, and members of the LGBTQIA+ community | Total calls: 40<br>Total emails: 83<br>Reasons for exclusion:<br>⟶ Participant's age did not match the criteria<br>⟶ Parents did not signed consent form<br>⟶ Low interest (stigma, coming out to the family, fear of bullying) |

**Supplementary Table 7. Mental health vignettes depicting prevalent conditions for each age group (using fictitious characters)**

| Condition | Groups | Vignette |
|---|---|---|
| Depression | Healthcare professionals<br>Teachers<br>NGO leaders<br>Caregivers of adolescents<br>Adolescents | Eleni is a 15-year-old who has been feeling unusually sad and miserable for the last few weeks. She is tired all the time and has trouble sleeping at night. Eleni doesn't feel like eating and has lost weight. She can't keep her mind on her activities or school, and her marks have dropped. She puts off making any decisions and even day-to-day tasks seem too much for her. |
| Conduct disorder | Healthcare professionals<br>Teachers<br>NGO leaders<br>Caregivers of 15 years old<br>15 years-old adolescents | Yannis is a 15-year-old who has been increasingly aggressive and intimidating around other people. Since elementary school, he has frequently initiated physical fights, offended other children, and threatened to hurt adults when bothered. In one of those fights, he and one of his classmates were hurt so badly that both of them had to be seen by a doctor. Once he set a fire in the schoolyard with his friends. He has been having difficulties following the school rules despite being punished by the School Principal. Yannis is explicitly lying to get out of trouble and stealing things from people around him. Yannis doesn't seem to be concerned about any of his behaviors, frequently stating that he doesn't care about anyone or anything. |
| Social anxiety | Eight year-old children and their caregivers | Eleni is an 8-year-old. Since starting her new school last year, she has become even more shy than usual and has made only one friend. She would really like to make more friends but is scared that she will do or say something embarrassing when she is around others. Although Eleni's work is OK, she rarely says a word in class and becomes incredibly nervous, trembles, blushes, and seems like she might vomit if she has to answer a question or speak in front of the class. With people she knows, Eleni is quite talkative, but she becomes quiet with anyone she doesn't know well. She never answers the phone and refuses to attend social gatherings. Her fears are unreasonable, but she can't seem to control them and this really upsets her. |
| ADHD | Eight year-old children and their caregivers | Yannis is an 8-year-old who has been feeling bored and agitated in the classroom. He has been talking a lot during classes, struggling to wait his turn in school activities, and leaving his seat without the teachers' permission. He finds it difficult to pay attention to details, avoids tasks that are considered difficult, and struggles to get schoolwork done. He is falling behind his classmates because he has been very distracted and has not been following instructions. Recently, he has been also fighting with some classmates and being rude to teachers. |

**Supplementary Table 8. Structure of the guiding manuals for focus groups**

| |
|---|
| **Introduction and ground rules** |
| **Views on well-being and mental health problems** |
| *All groups:*<br>Concepts of wellness<br>Concepts of mental health problem<br><br>*Only for caregivers, teachers, health professionals, and NGO leaders:*<br>How to notice and recognize if a child/adolescent is facing mental health problems?<br>When the child/adolescent needs additional help?<br>What are caregivers and professionals roles in that?<br>How would they approach the situation? |
| **Views on emotional problems** |
| Mental health vignette on **internalizing condition**<br>Views on what is happening to the child/adolescent (what cause, how common, have you encountered a situation like this?)<br>What should this person do and what would help<br>How could this person help themself of reach out for help and who to<br>Would this person feel comfortable in reaching out?<br>Opportunities for help in the community<br>What would do if facing/presencing someone facing a similar condition<br>The role of parents, teachers, and professionals |
| **Views on behavioral problems** |
| Mental health vignette on **externalizing condition**<br>Views on what is happening to the child/adolescent<br>What should this person do<br>How could this person help themself of reach out for help<br>Opportunities for help in the community<br>What would do if facing/presencing someone facing a similar condition<br>The role of parents, teachers, and professionals |
| **Stigma and discrimination** |
| What might keep people from talking about or seeking help?<br>What could help them out with feeling more comfortable in that? |
| **Services** |
| Places to reach in the community<br>Knowledge of where to reach<br>Availability and awareness of different mental health providers (mental health professionals, general health professionals, school staff, other professionals like religious leaders)<br>What kind of help available in each one<br>Obstacles in reaching them |

| **Access** |
| --- |
| Mental health care options and their perception of effectiveness<br>Understanding and attitudes towards modalities of treatment (psychotherapy, medication)<br>Barriers to access |
| **Training background, needs and perspectives** *(only for healthcare professionals, NGO leaders, and teachers)* |
| Challenges for the professional category when dealing with child and adolescent mental health<br>Resources in the institution they work with<br>Perceived skills<br>Which skills they want to acquire, if any<br>Perspective about training and formats<br>Obstacles for training |

**Supplementary Table 9. Extracts supporting focus groups' findings**

**<u>Children and Adolescents</u>**

- Children and adolescents believe mental health problems appear through actions and appearance, including rudeness, avoiding activities, or crying. However, some perceive such struggles may not be apparent to others or themselves.

⟶ ***"He/she may be shutting himself/herself in the room, with no appetite for anything, no mood to leave the house, eat, and with low performance in the classes."*** [12 to 18 year-old male]

⟶ ***"I think this person will be more antisocial than if he/she felt happy, without any appetite to do various activities."*** [12 to 18 year-old male]

- They note the role of external factors on mental health but acknowledge individual factors as well. They recognize vignettes of depression and ADHD, and even mention conditions like dyslexia.

⟶ ***"I have a friend who generally does not behave so well; she has some problems, does not behave well, has a bit of attention deficit in the class and she also has dyslexia."*** [6 to 12 year-old female]

- Stigmatized views are more disseminated among children and adolescents, such as discrediting disorders, believing people with depression should just feel better, and belittling symptoms.

⟶ ***"Such a person has no incentive to move on emotionally and mentally. It could be depression."*** [12 to 18 year-old male]

⟶ ***"Maybe Eleni is going through the stage of adolescence; I have friends in adolescence, I think it is not depression and because we all go through adolescence, we seem to exaggerate things on our minds to highlight that we have all kinds of problems and that we do not feel well."*** [12 to 18 year-old female]

- Peer stigma is a barrier to seeking help, as many would not want their peers to know they are seeing a professional. They believe seeing others using mental health services could help normalize care.

⟶ ***"I would not advise him to open up to his friends, but I would recommend he talked to his parents, about his problems and if he wants, I recommend going to a psychologist as well."*** [6 to 12 year-old female]

⟶ ***"I believe the main reason is that people are ashamed or afraid of being judged by others."*** [12 to 18 year-old male]

- Children and adolescents identify family and specialists as a source of help, and assume some problems need to be addressed by specialists.

⟶ ***"To speak to a teacher from her school, a psychologist ... and I think that when she speaks to a teacher, the latter will in turn talk to the headmaster about these problems and I think the school will be interested in this child since she doesn't feel well, and some measures will be taken. A psychologist will be called for the child's benefit."*** [12 to 18 year-old female]

⟶ ***"An expert could help her such as a psychologist. In addition, a friend whom she can open up to and ask for his help and advise her what to do. And her family being the closest she has."*** [12 to 18 year-old male]

⟶ ***"Obviously, if I felt like that, something should have happened. You don't feel like that out of the blue. If I thought it was important, I would speak to the most appropriate person, such as to a psychologist, an expert or if it were a family issue, something with my friends, my relations, or something like that, perhaps I would speak to my parents or my friends."*** [12 to 18 year-old female]

- Sometimes there is a disbelief that help could make them feel better or doubts they would be able to recognize help is needed.

⟶ ***"Because they think they will not be able to remedy it, to manage it; maybe they do not believe that the problem can be corrected..."*** [6 to 12 year-old female]

⟶ ***"It is this selfishness, egoism regarding how people will judge them; they think everything is well and rosy. They do not recognize their problem; they don't think there is anything to solve."*** [12 to 18 year-old male]

**Minority groups (LGBTQIA+, Pomak, Roma, and refugee adolescents)**

- LGBTQIA+ adolescents conceptualize that mental health problems may be experienced privately and not shown out.

⟶ ***"I will say something and I apologize if I trigger someone, but there exist suicidal trends, which is something that when you are at a strange stage in your psychology, at such a fragile age, it is something that is 'easy' to arise and I believe that while these people obviously need help, even if they do not cry it out themselves, if parents, if friends, if society learns something about them, they put them aside... if this person attempts what he/she wants to do and achieves it, everyone will say what a pity, why did he/she not speak up to get help? While when this person really needs their help, we marginalize it because we cannot handle it. And this is really sad..."*** [12 to 18 year-old LGBTQIA+ adolescent]

- When conceptualizing wellness, Pomak, refugees, and LGBTQIA+ adolescents referred to the importance of freedom and feeling free.

⟶ ***"At my age, it's to be happy, to be free, to be able to do whatever you want, to have no obstacles and to have some people with you. Or it could be friends or family or a sister/ brother…"*** [12 to 18 year-old Pomak adolescent]

⟶ ***"I think that a person who is happy will have a freedom to speak without fear of what will happen if he/she speaks."*** [12 to 18 year-old LGBTQIA+ adolescent]

- Some minority groups are especially vulnerable to stigmatized views. Refugee minors conceptualize mental health issues as “craziness” to be dealt with by deprivation in psychiatric hospitals. Pomak and Roma adolescents believe people with mental health problems should “distract” themselves and not seek professional assistance.

⟶ ***“One reason may be the stigma around asking for help on psychological issues… If you ask for help and if you go to a psychologist, let us say you are "crazy" or if you need some kind of professional help, this makes you weak or strange and all this stigma makes these people fearful, that if people around find out they have asked for help, they will not hang around with them, they will not talk to them, they will not be treated in the same way.”*** [12 to 18 year-old LGBTQIA+ adolescent]

⟶ ***“The best way is to go to the hospital, get well and come back.”*** [12 to 18 year-old refugee adolescent]

- Roma adolescents are less likely to seek assistance because they would not want to seek help outside of their community.

⟶ ***“Difficult… because I have my own people [Roma community members]; I would not go to them first.”*** [12 to 18 year-old Roma adolescent]

⟶ ***“I wouldn't address a specialist because I am an introvert person who does not open up easily and I would not feel comfortable speaking to a stranger, even if his job is to listen to people. I would prefer to talk to someone who is close to me. Unless the problem becomes bigger, and you will have to talk to the expert… Yes, I mean if my problem, if my psychological condition aggravated as time went by, I would go to a specialist…”*** [12 to 18 year-old Roma adolescent]

⟶ ***“One reason I wouldn't go to a specialist, it’s because you would not have the same trust as when talking to a well-known person.”*** [12 to 18 year-old Roma adolescent]

**Caregivers**

- Parents are aware of the importance of children and adolescents’ mental health, being willing to discuss and engage in learning about it.

⟶ ***“Conversations and meetings like this one ought to take place more often, maybe they should even be required in our workplace environments so that we can, as individuals, learn to better communicate with our families, our own personal environments, and be able to better understand those people with which we share those environments. I think it's really important to speak and feel that you're being heard, and to have ears to hear that extra thought that maybe you hadn't had, right?”*** [Parent of 6 to 12 year-old child]

- Parents are confident that they can identify mental health problems in their kids through signs such as behavior, uncontrolled emotions, shifts in routine, and social isolation.

⟶ ***“[Of] my three children, they have totally different reactions when they are not well. My son will shut himself off, in his room, more hours than before; we will call him to come to eat and he would say ‘I am not hungry, I will come later.’ There is generally a refusal to do things that he would otherwise do under different circumstances. One of my daughters will cling to the mobile phone or***

***the PC and will stay there for endless hours. She will isolate herself; my other daughter will be the nagging one. Just like a baby feeling asleep and wanting to go to sleep. Exactly this. She will be annoyed even with a mosquito flying from the outside. In other words, two of them are prone to isolation and the other one is nagging, grumbling and gets annoyed with everything ... Also, their look tells me things. I see the dark gaze when something goes wrong or when something has happened to them. But also, the way they look at each other, the way they look at me. It's a different look.”*** [Parent of 12 to 18 year-old adolescent]

- They are aware of the importance of the environment in mental health. Signs of mental health struggles are attributed to stressors such as suffering from love, isolation, fights, social media, or challenges at school.

⟶ ***“Well first of all you’ve got challenges children face in their immediate environment. Parents, grandparents. Then there's the school environment. Technology and how it's entered children's lives. That's all. I believe the challenges and stimuli are numerous, very numerous.”*** [Parent of 6 to 12 year-old child]

- Parents did not perform well in identifying mental health conditions when shown vignettes. Typical signs of ADHD were not recognized, having this manifestation attributed to issues at home or to the overuse of technology.

⟶ ***“It’s like in the first case with Eleni where I'd look into the school environment, here I think we're looking at a problem at home. This all originates with his family. Something's not right with Yannis, for him to display this sort of behavior towards his classmates and teachers, and for him not to be consistent in his classes. I'd look into that aspect of the family.”*** [Parent of 6 to 12 year-old child]

- There is a willingness to accept psychotherapy, but parents are reluctant towards prescription drugs.

⟶ ***“Sessions with a psychologist, a child psychologist. And, if needed, if the psychologist believes that psychiatric help is warranted, or pharmaceutical help maybe? But I'd start with sessions with a child psychologist.”*** [Parent of 6 to 12 year-old child]

⟶ ***“I would also be hesitant; if all other possibilities to solve the problem would have been exhausted, possibly I would consent to my child taking medication. And my reasoning behind it is not to pass a message to the child that for any problem we encounter in life, we take medication to solve it. To avoid it turning it into addiction. But it certainly depends on the case. For example, a child who is depressed needs medication to overcome it and you will not let it get worse and worse.”*** [Parent of 12 to 18 year-old adolescent]

- For parents, the initial step to addressing mental health issues is to try and solve it themselves or with advice from other parents, only reaching out to specialists if the issue persists.

⟶ ***“I simply think that if my child would have an issue, I would try to search a little on my own at the beginning or I would talk to other moms with whom I have contacts and we talk to each other, but I also think it is very important to address a specialist. In other words, I think that this issue is taboo today and we are not talking about it; most of the people are dealing with such an issue and they don’t speak up, because they may be ashamed, or they may think that they will be judged by others or anything. Well, but I think it is very important and essential to address a specialist.”*** [Parent of 12 to 18 year-old adolescent]

- Parents recognize that taboo may be a barrier to seek for help, but none recognize it as a barrier for themselves.

⟶ ***“I think it's still considered a taboo topic in Greece, that somebody may have a mental health issue that requires the attention of a professional. He's still considered the “quack”, you know, an expression used for that [...] they’ll stick a label on you and, well, you’ll be stuck with it for the rest of your life. It’s still a taboo topic, we aren't yet comfortable with approaching mental health professionals and being okay with it. Even those who do in fact seek them out, I think. They wouldn't open up about it.”*** [Parent of 6 to 12 year-old child]

- Several parents spoke of their personal lack of understanding and struggles opening up about mental health.

⟶ ***“Here’s what scares me. First of all, we have to find the reason why Eleni is experiencing what she is. Because in order to help her, we have to find out what's wrong. If, for example, it has its origins in her home environment, how can her parents help her when it's them who’ve caused the problem if they've caused it? When she goes to school, how can she be helped if they don't know about it at home? So, it's a vicious cycle and it all scares me to think about how a child can find help if you don't know... Obviously, you need a specialist. You need professional help. But I think we need to start from the fact that the first person to understand that something's wrong with Eleni ought to speak with a professional to start to untie the knot, to help her. And for those who've caused the problem to find help as well.”*** [Parent of 6 to 12 year-old child]

**Teachers and Healthcare Professionals**

- There is a positive attitude towards mental health among all professional categories, who feel responsible for caring for children and adolescents.

⟶ ***“I believe in the idea that mental health is as important as physical health and that in the same way they would go to the doctor for some physical problem, so they should also do the same and if they have some psychological problem.”*** [Teacher of 12-16 years-old Students]

- They understand that environmental factors are determinants of mental health, including violence at home, parental neglect, or school problems. They are also aware of specific struggles faced by minorities, including race, sexuality, and disability.

⟶ ***“I would also use the word inclusion, not only in relation to children, refugees and migrants, but I would include all children in adolescence; the need for inclusion is a challenge for them, and I think that this affects them. I also think that the demands or the expectations of the family environment are issues representing a challenge for adolescents and I would certainly add the pandemic and what comes after it. In other words, I think that we are now in a completely different phase; in particular, the period of pandemic and lockdown has had a very great influence on the children who were in adolescence and as it appears, we will witness similar situations soon.”*** [Non-Governmental Organization Worker]

⟶ ***“Look, one of the many challenges refers to the new subjects which we face. Migration, violence, abuse. That is, crises. I think that we are intercultural, which is all right, and at the same time identical to immigration issues. We take care of many new things. things... gender dysphoria, so we have many new demands, new kinds of demands, new ones, young people, people who are suffering, facing new challenges, so all this probably goes to the need for continuing education, let us say so, which is one of the closing issues. A change in the subject generates changes in***

***pathologies, that is, an update, an update of the answers, the subject matter."*** [Child Psychiatrist]

- Healthcare providers see both parents and children struggling to understand the different facets of mental health, with limited awareness of the available resources. They believe more information for the general public is crucial.

⟶ ***"That it's not clear enough what mental health means. I mean, we treat it as a taboo and children may be afraid to communicate about their condition, they may confuse a psychiatric condition with a psychological one and the party who will prescribe medication, if they need to take any; I think it is the psychiatrist who can prescribe medication and not the psychologist, to whom we refer all this time. So we don't know exactly what help means and in what context it will be provided and what communication with a psychologist and a psychiatrist means. Generally, the context is not very clear. Like visiting a pathologist or some other specialty would be."*** [Teacher of 12-16 years-old Students]

⟶ ***"The truth is that we have not been trained to approach all mental health issues, whether we are the person being involved or the person on the opposite side. So, I think that education for everyone, and for health professionals [is needed], because we cannot easily approach such an issue either. But on the other hand, you need training also to externalize your emotions. Training is needed, with whatever this entails. The truth is that our educational system has not adopted such an approach."*** [Primary Health Care Nurse]

- Health professionals see stigma as a problem in Greece, especially among adults. The discomfort when talking about mental health would hold people from seeking help. This is especially pronounced in remote areas, with rural inhabitants even looking for assistance in urban areas as they worry about being perceived as weak.

⟶ ***"You know, sometimes we say about the stigma of mental disorders, that "it's old news", "it's gone", "it's outdated", "it's a mentality we've left behind us" ... but that's not the case. Especially I don't want to say this either, but I will. Especially in smaller societies, things are even harder about this matter. There are many thing holding people back. Holding them back from the next step."*** [Social Worker]

- Teachers note that peer and school community acceptance or stigmatization is a challenge and point out that students fear friends will find out they are seeking assistance.

⟶ ***"Yes, I certainly agree there's this taboo among teenagers perhaps before when they tell jokes with each other; so, they are not willing to speak up about their problems, what concerns them to avoid being scorned or made a fool of by their peers. What needs to be done is essentially to provide as safe an environment as possible for young people to be able to open up to the experts when there is [...] to be able to start the treatment immediately."*** [Non-Governmental Organization Worker]

⟶ ***"The attitude of their classmates, if they are not open to this process and do not support them and they can stigmatize children, who face such problems. How the school community itself accepts and embraces children facing such issues."*** [Teacher of 12-16 years-old students]

- Teachers and health professionals stress that one of the biggest challenges is overcoming parental refusal that care is needed. Health providers attribute it to not knowing treatment options and fearing what they may entail.

⟶ ***“Unfortunately, it is the refusal of parents to accept that there is a problem. The parent's refusal to engage in a fruitful dialogue. The parent's refusal to refer to a specialist. This is what I encounter much more often.”*** [Primary Health Care Nurse]

⟶ ***“Parents really do not accept the problems that may be created, either within the family environment or within the school environment. In specific, I'm going to talk about my own school environment and tell you that I play the role of the teacher and of the supervisor and of the psychologist and the janitor... I play all roles. I’m wearing many hats. Returning to the previous example I gave you with the little girl the mother did not want to follow the school procedure and seek the help of an expert, as she was concerned that this would stigmatize the child and that it would become known in this small, closed society where the school is located. So, we also tried to figure out a solution between us. The mother, the child, me and probably because she asked me, the mother of the other children. In other words, this year I have to say that I was faced with a difficult situation, and I did not know how to handle it many times. On the one hand, I had the mother, who was insistent on finding a solution and wanted me to intervene and talk to the mums of the other children and on the other hand, she would not accept to move forward and get the opinion of an expert.”*** [Teacher of 8-14 years-old students]

- Healthcare professionals believe it is necessary to improve relationships between caregivers and children/adolescents so that they feel more comfortable expressing their struggles.

⟶ ***“I don't want to say anything about pharmaceutical treatment, but I do want to say that psychotherapy can't be on its own. I mean, I don't know how [Redacted] can manage; she only sees kids and teenagers, but in my mind, this can't exist on its own. There must also be something that covers the people of the close environment. Another colleague who does parallel work with the family.”*** [Female Social Worker]

⟶ ***“To include family management, too. This is not for granted. We are talking only about the child, fine, to have tools, but also about the management of caregivers. Training, management approach and communication capability of the problem.”*** [Primary Health Care Nurse]
